# Comparison of Machine Learning Algorithms for the Prediction of Current Procedural Terminology (CPT) Codes from Pathology Reports

**DOI:** 10.1101/2021.03.13.21253502

**Authors:** Joshua Levy, Nishitha Vattikonda, Christian Haudenschild, Brock Christensen, Louis Vaickus

## Abstract

**Background:** Pathology reports serve as an auditable trail of a patient’s clinical narrative containing important free text pertaining to diagnosis, prognosis and specimen processing. Recent works have utilized sophisticated natural language processing (NLP) pipelines which include rule-based or machine learning analytics to uncover patterns from text to inform clinical endpoints and biomarker information. While deep learning methods have come to the forefront of NLP, there have been limited comparisons with the performance of other machine learning methods in extracting key insights for prediction of medical procedure information (Current Procedural Terminology; CPT codes), that informs insurance claims, medical research, and healthcare policy and utilization. Additionally, the utility of combining and ranking information from multiple report subfields as compared to exclusively using the diagnostic field for the prediction of CPT codes and signing pathologist remains unclear.

**Methods:** After passing pathology reports through a preprocessing pipeline, we utilized advanced topic modeling techniques such as UMAP and LDA to identify topics with diagnostic relevance in order to characterize a cohort of 93,039 pathology reports at the Dartmouth-Hitchcock Department of Pathology and Laboratory Medicine (DPLM). We separately compared XGBoost, SVM, and BERT methodologies for prediction of 38 different CPT codes using 5-fold cross validation, using both the diagnostic text only as well as text from all subfields. We performed similar analyses for characterizing text from a group of the twenty pathologists with the most pathology report sign-outs. Finally, we interpreted report and cohort level important words using TF-IDF, Shapley Additive Explanations (SHAP), attention, and integrated gradients.

**Results:** We identified 10 topics for both the diagnostic-only and all-fields text, which pertained to diagnostic and procedural information respectively. The topics were associated with select CPT codes, pathologists and report clusters. Operating on the diagnostic text alone, XGBoost performed similarly to BERT for prediction of CPT codes. When utilizing all report subfields, XGBoost outperformed BERT for prediction of CPT codes, though XGBoost and BERT performed similarly for prediction of signing pathologist. Both XGBoost and BERT outperformed SVM. Utilizing additional subfields of the pathology report increased prediction accuracy for the CPT code and pathologist classification tasks. Misclassification of pathologist was largely subspecialty related. We identified text that is CPT and pathologist specific.

**Conclusions:** Our approach generated CPT code predictions with an accuracy higher than that reported in previous literature. While diagnostic text is an important information source for NLP pipelines in pathology, additional insights may be extracted from other report subfields. Although deep learning approaches did not outperform XGBoost approaches, they may lend valuable information to pipelines that combine image, text and -omics information. Future resource-saving opportunities exist for utilizing pathology reports to help hospitals detect mis-billing and estimate productivity metrics that pertain to pathologist compensation (RVU’s).

## Background and Significance

Electronic Health Records (EHR) ^1^ refers to both the structured and unstructured components of patients’ health records/information (PHI), synthesized from a myriad of data sources and modalities. Such data, particularly clinical text reports, are increasingly relevant to “Big Data” in the biomedical domain. While structured components of EHR, such as clinical procedural and diagnostic codes, are able to effectively store the patient’s history ^2–4^, unstructured clinical notes reflect an amalgamation of more nuanced clinical narratives. Such documentation may serve to refresh the clinician on the patient’s history, highlight key aspects of the patient’s health, and facilitate patient handoff among providers. Furthermore, analysis of clinical free text may reveal physician bias or inform an audit trail of the patient’s clinical outcomes for purposes of quality improvement. As such, utilizing sophisticated algorithmic techniques to assess text data in pathology reports may improve decision making and hospital processes/efficiency, possibly saving hospital resources while prioritizing patient health.

Natural language processing (NLP)^3,5–8^, is an analytic technique to extract semantic and syntactic information from textual data. Traditionally, rule-based approaches cross-reference and tabulate domain-specific key words or phrases with large biomedical ontologies and standardized vocabularies such as Unified Medical Language System (UMLS)^9,10^. However, although these approaches provide accurate means of assessing a narrow range of specified patterns, they are neither flexible nor generalizable since they require extensive annotation and development from a specialist. Machine learning approaches (e.g. support vector machines, random forest) ^11,12^, employ a set of computational heuristics to circumvent manual specification of search criteria to reveal patterns and trends in the data. While bag-of-word approaches^13,14^ study the frequency counts of words (unigrams) and phrases (bigrams, etc.) to compare the content of multiple documents for recurrent themes, deep learning approaches^15–17^ simultaneously capture syntax and semantics with artificial neural network techniques. Recent deep learning NLP approaches have demonstrated the ability to capture meaningful nuances that are lost in frequency-based approaches; for instance, these approaches can effectively contextualize short- and long-range dependencies between words^18,19^. Despite potential advantages conferred from less structured approaches, analysis of text across any domain usually necessitates balancing domain-specific customization (e.g. a medical term/abbreviation corpora) with generalized NLP techniques.

Analysis of pathology reports using NLP has been particularly impactful in recent years, particularly in the areas of information extraction, summarization, and categorization. Noteworthy developments include information extraction pipelines which utilize regular expressions (regex), to highlight key report findings (eg. extraction of molecular test results) ^20– 23^, as well as topic modeling approaches that summarize a document corpus by common themes and wording^24^. In addition to extraction methods, machine learning techniques have been applied to classify pathologist reports^25^; notable examples include prediction of ICD-O morphological diagnostic codes ^26,27^ and prediction of CPT codes based only on diagnostic text ^28,29^. Widespread misspelling of words and jargon specific to individual physicians have made it difficult to reliably utilize the rule-based and even machine learning approaches for report prediction in a clinical workflow. Additionally, hedging and uncertainty in text reports may further obfuscate findings^30^.

## Objective

In this study, we sought to predict the assignment of 38 different CPT procedure codes across a large corpus of over 93,039 pathology reports using XGBoost, Support Vector Machine (SVM) and BERT (Bidirectional Encoder Representation from Transformers) techniques from the Dartmouth-Hitchcock Department of Pathology and Laboratory Medicine (DPLM), a mid-sized academic medical center. Furthermore, we explored methods that incorporate, into the deep learning modeling approach, other document subfields outside of the diagnostic text, which may contain additional information. Finally, we utilized these technologies to investigate physician-specific or uncertain word choices that could have biased results.

## Approach and Procedure

### Data Acquisition

We obtained an Institutional Review Board approval and accessed over 96,418 pathologist reports from DPLM, collected between June 2015 and June 2020. We removed a total of 3,379 reports that did not contain any diagnostic text associated with CPT codes, retaining 93,039 reports (**Supplementary Table 1**). Each report was appended with metadata including corresponding EPIC (EPIC systems, Verona, WI)^31^, Charge Description Master (CDM), and CPT procedural codes, the practicing pathologist, the amount of time to sign out the document, and other details. A fuzzy word matching system was deployed to resolve misspellings between documents. The documents were deidentified by stripping all PHI-containing fields and numerals from the text and replacing with holder characters (e.g. 87560 becomes #####).A publicly available database of hundreds of thousands of first and last names was utilized to remove all mention of names in the report text as a final check.

### Preprocessing

The text was preprocessed using the Spacy package^32^, to tokenize text and removed stop words. We also split up each pathology report into their structured sections: Diagnosis, Clinical Information, Specimen Processing, Discussion, Additional Studies, Results, Interpretation. This allowed for an equal comparison between the machine learning algorithms. The deep learning algorithm BERT can only operate on 512 words at a time due to computational constraints.

Sometimes, the pathology reports exceeded this length when considering the entire document (1.77% exceeded 512 words) and as such these reports were limited to the diagnosis section (0.02% exceeded 512 words) when training a new BERT model (**Supplementary Table 1; Supplementary Figure 1**). We removed all pathology reports which did not contain a diagnosis section.

### Characterization of the Text Corpus

After preprocessing, we encoded each report tabulating the occurrence of all contiguous one-to two-word sequences (unigram and bigrams) to form sparse count matrices, where each column represents a word or phrase and each row represents the document, and the value is the frequency of occurrence in the document. While the term frequency may be representative of the distribution of words/phrases in a corpus, high frequency words that are featured across most of the document corpus are less likely to yield informative lexicon that is specific to a subset of the documents. To account for less important but ubiquitous words, we transformed raw word frequencies to term frequency inverse document frequency (tf-idf) values, which up-weights the importance of the word based on its occurrence within a specific document (term frequency), but down-weights the importance if the word is featured across the corpus (inverse document frequency) (see Supplementary Material, section “Additional Description of Topic Modeling and Report Characterization Techniques”). We summed the tf-idf value of each word across the documents to capture the word’s overall importance across the reports and utilized a word cloud algorithm to display the relative importance of the top words.

After constructing count matrices, we sought to characterize and cluster pathology documents as they relate to each other and ascribe themes to the clusters. UMAP ^33^ dimensionality reduction was used to project the higher dimensional word frequency data into lower dimensions while preserving important functional relationships. Each document could then be represented by a 3D point in the Cartesian coordinate system; these points were clustered using a density-based clustering algorithm called HDBSCAN ^34^ to simultaneously estimate characteristic groupings of documents while filtering out noisy documents which did not explicitly fit in these larger clusters. To understand which topics were generally present in each cluster we deployed Latent Dirichlet Allocation (LDA)^13^, which identifies topics characterized by a set of words, then derives the distribution of topics over all clusters. This is accomplished via a generative model which attempts to recapitulate the original count matrix, further outlined in greater detail in the Supplementary Material, section “Additional Description of Topic Modeling and Report Characterization Techniques”. The individual topics estimated using LDA may be conceptualized as a Dirichlet/multinomial distribution (“weight” per each word/phrase) over all unigrams and bigrams, where a higher weight indicates membership in the topic. The characteristic words pertaining to each topic were visualized using a word cloud algorithm. Finally, we correlated the CPT codes with clusters, topics, and select pathologists using Point-Biserial and Spearman correlation measures ^35^ to further characterize the overall cohort.

### Machine Learning Models

We implemented the following three machine learning algorithms in our study as a basis for our text classification pipeline (**Figure 1**):

**Figure 1:**
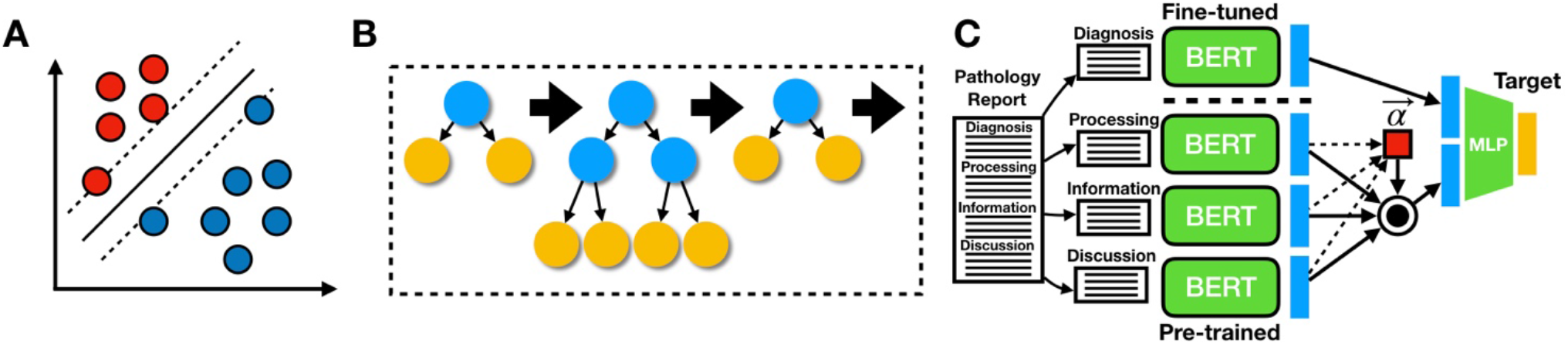
Model Descriptions: Graphics depicting: **A)** SVM, where hyperplane linearly separates pathology reports, which are represented by individual datapoints; **B)** XGBoost, which sequentially fits decision trees based on residuals from sum of conditional means of previous trees and outcomes; **C)** *All-Fields* BERT model, where a diagnosis-specific neural network extracts relevant features from the diagnostic field, while a neural network trained on a separate clinical corpus extracts features for the remaining subfields; subfields are weighted and summed via the attention mechanism, indicated in red; subfields are combined with diagnostic features and finetuned with a multi-layer perceptron for the final prediction

#### SVM

We trained a Support Vector Machine model (SVM) ^36,37^ to make predictions using the UMAP embeddings formed from the tf-idf matrix. SVM operates by learning a hyperplane that obtains maximal distance (margin) to datapoints of a particular class (**Figure 1A**). However, because datapoints/texts from different classes may not be separable in the original embedding space, the SVM model projects data to a higher dimensional space where data can be linearly separated. We utilized GPU resources via the ThunderSVM package ^38^ to train the model in reasonable compute time.

#### Bag of Words with XGBoost

XGBoost algorithms ^39^ operate on the entire word by report count matrix and ensemble or average predictions across individual Classification and Regression Tree (CART) models ^40^. Individual CART models devise splitting rules that partition instances of the pathology notes based on whether the count of a particular word or phrase in a pathology note exceeds an algorithmically derived threshold. Important words and thresholds (i.e. partition rules) are selected from the corpus based on their ability to partition the data, based on the purity of a decision leaf through calculation of an entropy measure. Each successive splitting rule serves to further minimize the entropy or maximize the information gained. While Random Forest models ^41^ bootstrap which subsets of predictors/words and samples are selected for a given splitting rule of individual trees and aggregate the predictions from many such trees, Extreme Gradient Boosting Trees (XGBoost) fits trees (structure and the conditional means of the terminal nodes) sequentially based on the residual (in the binary classification setting, misclassification is estimated using a Bernoulli likelihood) between the outcome and the sum of bot conditional means of the previous trees (which are set) and the conditional mean of the current tree (which is optimized). This gradient-based optimization technique prioritizes samples with a large residual/gradient from the previous model fit to account for the previous “weak learners” (**Figure 1B**). In both scenarios, random forest (a bagging technique) and XGBoost (a boosting technique), individual trees may exhibit bias but together cover a larger predictor space. Our XGBoost classifier models were trained using the XGBoost library, which utilizes GPUs to speed up calculation.

#### BERT

Artificial neural networks (ANN) ^42^ are a class of algorithms that use highly interconnected computational nodes to capture relationships between predictors in complex data. The information is passed from the nodes of an input layer to individual nodes of subsequent layers that capture additional interactions and nonlinearities between predictors while forming abstractions of the data in the form of intermediate embeddings. The BERT (Bidirectional Encoder Representations from Transformers) ^18^ model first maps each word in a sentence to its own embedding and positional vectors, which captures key semantic/syntactic and contextual information that is largely absent from the Bag of Words approaches. These word level embeddings are passed to a series of self-attention layers (the Transformer component of the BERT model) which contextualizes the information of a single word in a sentence based on short- and long-term dependencies between all words from the sentence. The individual word embeddings are combined with the positional/contextual information, obtained via the self-attention mechanism, to create embeddings that represent the totality of a sentence. Finally, this information is passed to a series of fully connected layers that produce the final classification. With BERT, we are also able to analyze the relative importance and dependency between words in a document by extracting “attention matrices”. We are also able to retrieve sentence level embeddings encoded by the network by extracting vectors from the intermediate layers before it passes to final classification.

We trained the BERT models using the *HuggingFace Transformers* package ^43^, which utilizes GPU resources through the PyTorch framework. We used a collection of models that have already been pretrained on a large medical corpus ^44^ in order to both improve the predictive accuracy of our model and significantly reduce the computational load compared to training a model from scratch. Because significant compute resources are still required to train the model, most BERT models limit the document characterization length to 512 words. To address this, we split pathology reports into document subsections when training BERT models.

In training a BERT model, we updated the word embeddings through finetuning a pretrained model on our diagnostic corpus. This model, which had been trained solely on diagnostic text, could be used to predict the target of interest (*Dx Model*). However, we then used this fine-tuned model to extracted embeddings that were specific to the diagnosis subfield to serve as input for a model that could utilize text from other document subfields. We separately utilized the original pretrained model to extract embeddings from the other report subfields which are less biased by diagnostic code and thus more likely to provide contextual information (*All Fields Model*). We developed a global/gating attention mechanism procedure that serves to dynamically prune unimportant, missing, or low-quality document subsections for classification (**Figure 1C**).

Predictions may be obtained when some/all report subfields are supplied via the following method:

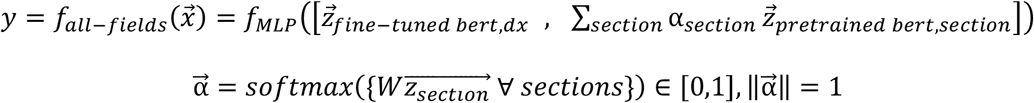

Where 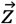 represents embeddings extracted from the pretrained and fine-tuned BERT embeddings on respective report subsections and 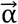 is a vector of attention scores between 0 and 1 that dictates the importance of particular subsections. Finally, *W* is a 768-dimension (dimensionality of BERT embeddings) by 1-dimensional matrix that generates the attention scores, while *f*_*MLP*_ are a set of fully connected layers that operate on the concatenation between the BERT embeddings that were finetuned on the diagnosis-specific section and those extracted using the pre-trained BERT model on the other document subfields.

### Prediction of CPT Codes

Using these machine learning techniques, we sought to predict each of the 38 different CPT codes (38 codes remained after removing codes that occurred less than 150 times across all sign-outs) using BERT, XGBoost and SVM. Given the characterization of the aforementioned deep learning framework, we utilized a BERT model that was pretrained first on a large corpus of biomedical research articles from PubMed, then pretrained using a medical corpus of free text notes from an intensive care unit (MIMIC3 database; Clinical-BioBERT) ^44–46^. Finally, the model was fine-tuned on our DHMC pathology report corpus (to capture institution-specific idiosyncrasies) for the task of classifying particular CPT codes from diagnostic text. XGBoost was trained on the original count matrix, while SVM was trained on a 6-dimensional UMAP projection; a UMAP projection was utilized for computational considerations. The models were evaluated using 5-fold cross validation for each CPT code. For each approach, we separately fit a model considering only the Diagnosis text (*Dx* Models) and all of the text (*All Fields* Models) to provide additional contextual information. We calculated the Area Under the Receiver Operating Curve (AUC-Score; considers sensitivity/specificity of the model at a variety of probability cutoffs; anything above a 0.5 AUC is better than random) for each CPT code to further explore reasons that some codes yielded lower scores than others. We also compared different algorithms via the sensitivity/specificity reported via their Youden’s index (the optimal tradeoff possible between sensitivity/specificity from the receiver operating curve), averaged across validation folds. Finally, we used Shapley Additive Explanations (SHAP; a model interpretation technique that estimates contributions of predictors to the prediction through credit allocation) ^47^ to estimate which words were important for classification of each of these codes, visualized using a word cloud. For the BERT model, we utilized the Captum ^48^ framework to visualize backpropagation from the outcome to predictors/words via IntegratedGradients ^49^ and attention matrices. Additional extraction of attention weights also revealed not only which words and their relationships contributed to prediction of the CPT code (i.e. self-attention denotes word-to-word relationships), but also which document subfields other than the diagnosis field were important for assignment of procedure code (i.e. global/gating attention prunes document subfields by learning to ignore irrelevant information; the degree of pruning can be extracted during inference). Further description of these model interpretability techniques (SHAP, Integrated Gradients, Self-Attention / “word-to-word”, Attention) may be found in the supplementary material (section “Additional Description of Explanation Techniques: SHAP, Integrated Gradients, Self-Attention, Attention Over Pathology Report Subfields”).

### Predictions of Physician Specific Language

We similarly trained all models to recognize the texts of the twenty pathologists with the most sign-outs to see whether the models could reveal pathologist-specific text. We retained reports from the twenty pathologists with the most sign-outs, reducing our document corpus from 93,039 documents to 64,583 documents, and utilized all three classification techniques to predict each sign-out pathologist simultaneously. The selected pathologists represented a variety of specialties. Choosing only the most prolific pathologists allowed us to test our hypothesis of pathologist-specific language without being subject to spurious associations by a rare outcome in the multi-class setting. Pathologist specific word choice was extracted using SHAP/Captum from the resulting model fit and visualized using word clouds and attention matrices.

## Results

### Corpus Preprocessing and UMAP Results

After initial filtering, we amassed a total of 93,039 pathology reports, which were broken into the following subsections: Diagnosis, Clinical Information, Specimen Processing, Discussion, Additional Studies, Results, and Interpretation. The median word length per document was 119 words (Interquartile Range; IQR=90). Very few reports contained subfields that exceeded the length acceptable by the BERT algorithm (2% of reports containing a *Results* section exceeded this threshold; **Supplementary Table 1; Supplementary Figure 1**).

Displayed first are word clouds of the top twenty-five words in only the diagnostic document subsection (**Figure 2A**) and across all document subsections (**Figure 2B**), with their size reflecting their tf-idf scores (**Figure 2A-B**). As expected, the diagnostic-field cloud contains words that are pertinent to the main diagnosis, while the all-field cloud contains words that are more procedural, suggesting that other pathology document subfields yield distinct and specific clinical information that may lend complementary information versus analysis solely on diagnostic fields. We clustered and visualized the diagnostic subsection and also all document subsections after running UMAP, which yielded 8 and 15 distinct clusters respectively (**Figure 2C-D**). Number of words per report correlated poorly with the number of total procedural codes assigned (spearman *r = 0*.*066,p <0*.*01*). However, when these correlations were assessed within the HDBSCAN report clusters (subset to reports within a particular cluster for cluster-specific trends), 33% of the all-fields report clusters reported moderate correlations (**Supplementary Table 2**). Interestingly, one of the eight report clusters from the diagnostic fields experienced a moderate negative correlation with number of codes assigned.

**Figure 2:**
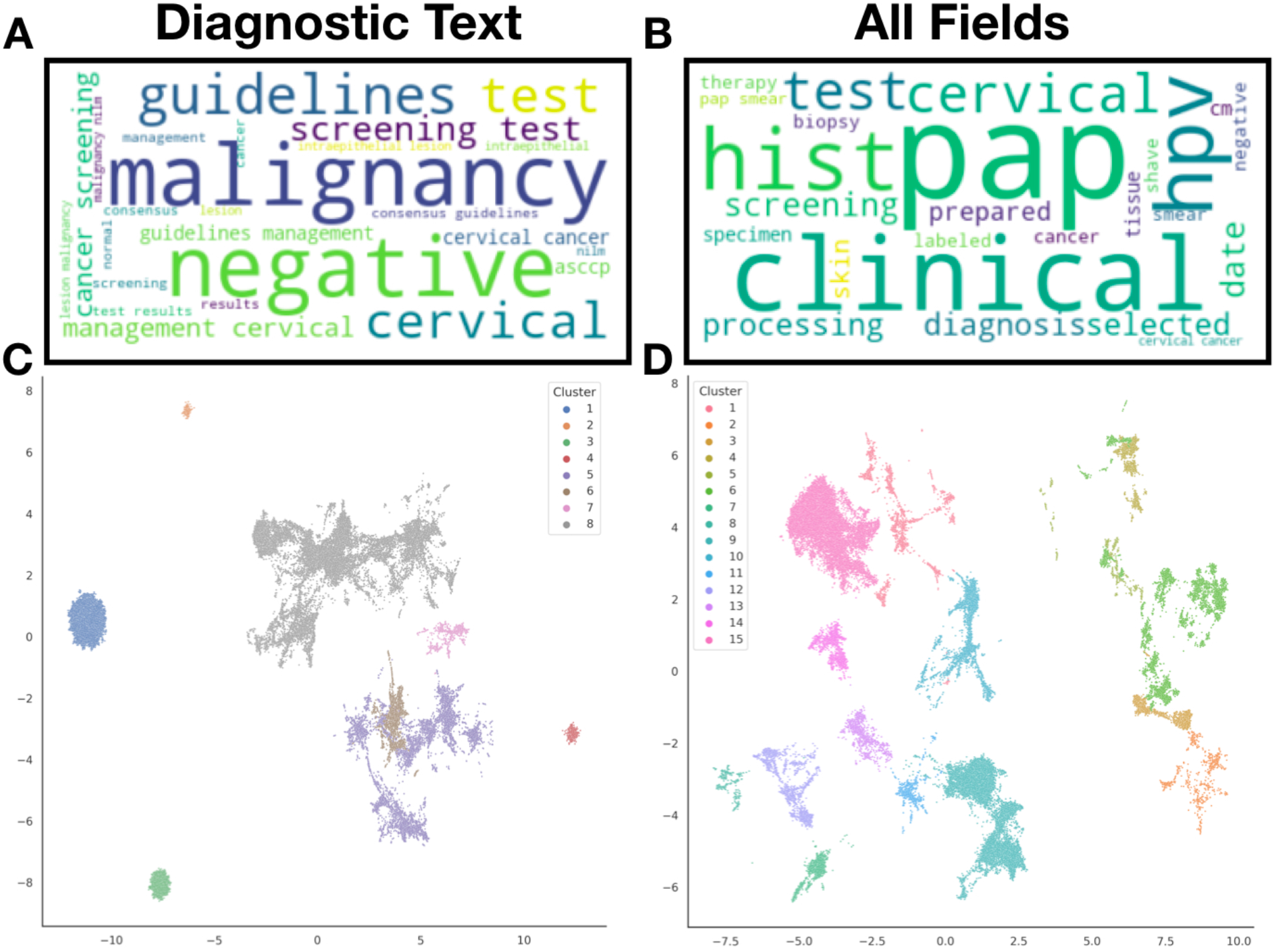
Pathology report corpus characterization: **A-B)** Word cloud depicting words with highest aggregated tf-idf scores across the corpus of: **A)** diagnostic text only, **B)** all report subfields (*all-fields*); important words across the corpus indicated by relative size of the word in the word cloud; **C-D)** UMAP projection of the tf-idf matrix, clustered and noise removal via HDBSCAN for: **C)** diagnostic texts only, **D)** all report subfields (*all-fields*)

### Topic Modeling with LDA and Additional Topic Associations

From our LDA analysis on all document subsections, we discovered 10 topics (**Figure 3; Supplementary Table 3**). Correlations between these topics with clusters, pathologists, and CPT codes are displayed in the supplementary material (**Supplementary Figures 2-4**). We discovered additional associations between CPT codes, clusters and pathologists (**Supplementary Table 5, Supplementary Figure 6A**), suggesting a specialty bias in document characterization. We clustered pathologists using co-occurrence of procedural code assignments in order to establish “subspecialties” (eg. pathologist who signs out multiple specialties) which could be used to help interpret sources of bias in evaluation of downstream modeling approaches.

**Figure 3:**
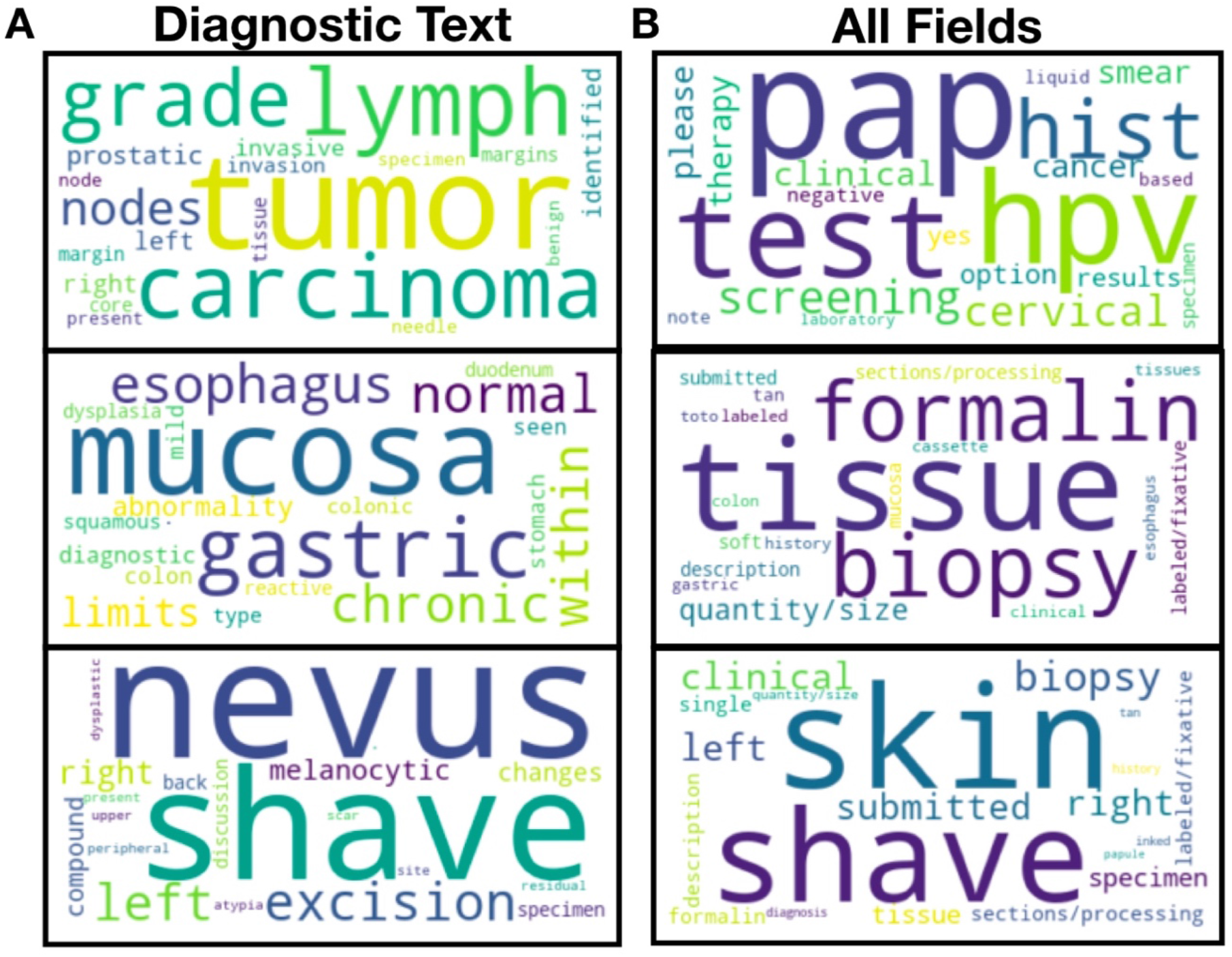
LDA Topic Words: Important words found for three select LDA Topics from: **A)** diagnostic text only, **B)** all report subfields (*all-fields*); important words across the corpus indicated by relative size of the word in the word cloud

### CPT Code Classification

We were able to accurately assign a CPT code to each document, regardless of which machine learning algorithm was utilized (**Table 1; Figure 4A; Supplementary Table 4**). We found that XGBoost (median AUC=0.985) performed comparably to BERT (median AUC=0.990; p=0.64) when predicting CPT codes based on the diagnostic subfield alone, while SVM performed worse (median AUC=0.990) than both approaches, per cross-validated AUC statistics (**Table 1; Figure 4A; Supplementary Tables 4-5; Supplementary Figure 7**). We also discovered that classifying by including all of the document sub-elements performed better than just classifying based on the diagnostic subsection (p<0.001 for both BERT and XGBoost approaches; **Supplementary Table 5; Supplementary Figure 7**), suggesting that these other more procedural / descriptive elements contribute meaningful contextual information for the assignment of CPT codes. XGBoost (median AUC=0.997) outperformed BERT (median AUC=0.995) statistically (p<0.001) when utilizing all of the report subfields but given the high predictive performance these differences were not meaningful. Plots and tabulated statistics of the Youden Index derived from sensitivity/specificity of these algorithms across all of the validation folds confirm that utilizing information from all report subfields is better than utilizing information from the diagnostic text (**Supplementary Table 6; Supplementary Figure 8**). Averaging Youden’s J statistic across all XGBoost and deep learning models, codes for immunohistochemistry/cytochemistry (CPT 88341, 88342, 88344, 88360), surgical pathology (CPT 88305) and flow cytometry (CPT 88188, 88189) performed worse versus other procedural codes; however, the performance improved considerably when including all report subfields for these codes (**Supplementary Tables 6-7**). Interestingly, the code for cytogenetic testing (CPT 88271) also experienced large improvements in sensitivity and specificity by incorporating other report subfields (**Supplementary Table 7**).

**Table 1:**
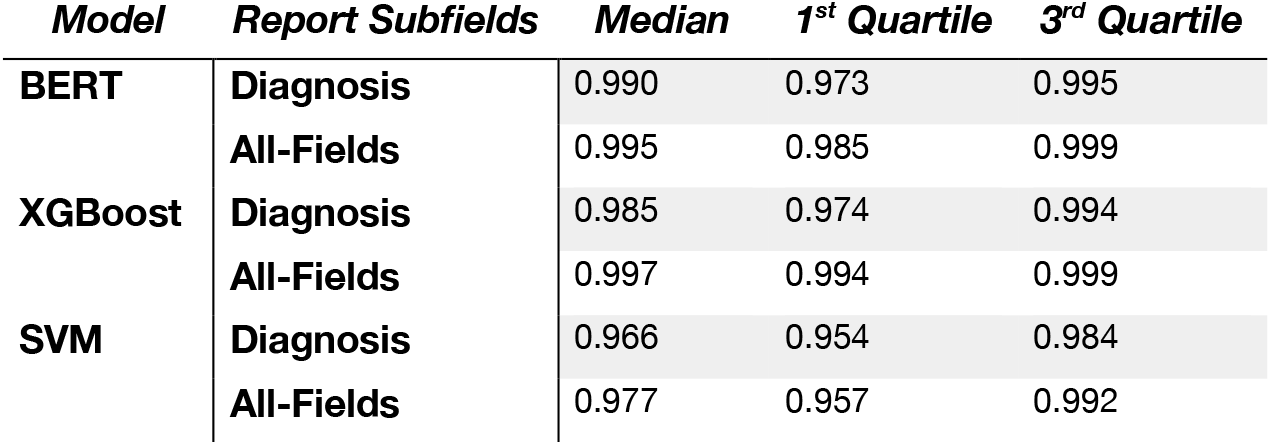
Summary of distribution of AUCs across CPT codes for BERT, XGBoost and SVM prediction models for diagnostic and *all-fields* text

| *Model* | *Report Subfields* | *Median* | *1<sup>st</sup> Quartile* | *3<sup>rd</sup> Quartile* |
| --- | --- | --- | --- | --- |
| **BERT** | **Diagnosis** | 0.990 | 0.973 | 0.995 |
|  | **All-Fields** | 0.995 | 0.985 | 0.999 |
| **XGBoost** | **Diagnosis** | 0.985 | 0.974 | 0.994 |
|  | **All-Fields** | 0.997 | 0.994 | 0.999 |
| **SVM** | **Diagnosis** | 0.966 | 0.954 | 0.984 |
|  | **All-Fields** | 0.977 | 0.957 | 0.992 |

**Figure 4:**
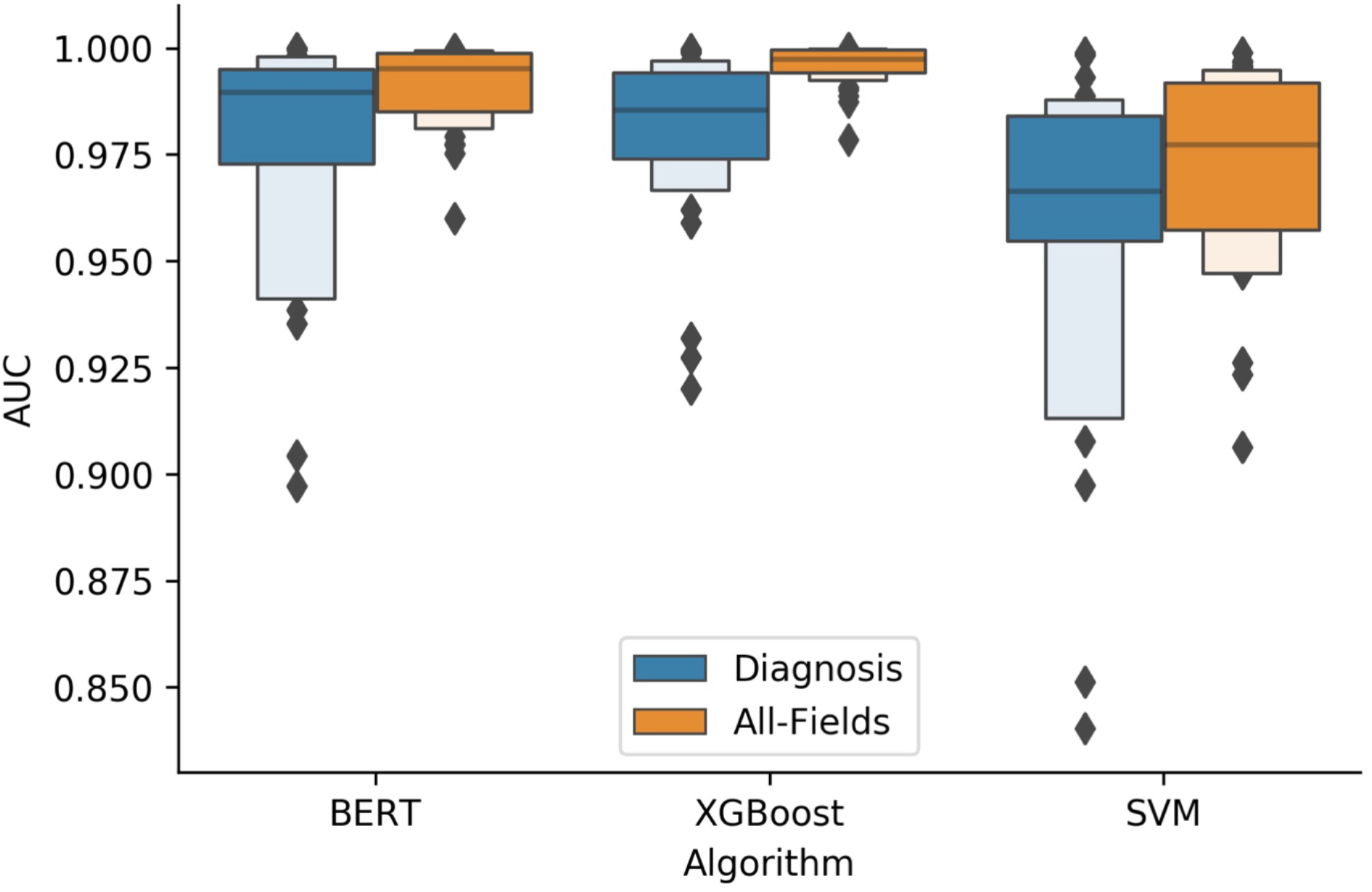
CPT Code Model Performance: Grouped boxenplots demonstrating performance of machine learning models (BERT, XGBoost, SVM) across CPT codes (distribution of AUCs reported for each CPT code), given analysis of either the diagnostic text (blue) or all report subfields (orange)

We also visualized which words were found to be important for a subsample of diagnostic codes using the XGBoost algorithm (**Figure 5**). Reports that were assigned the same CPT code clustered together in select low dimensional representations learned by some of the *All Fields* BERT models (**Figure 6A,C,E**). Model-based interpretations of a few sample sentences for CPT codes using the *Diagnosis* BERT approach revealed important phrases that aligned with assignment of the respective CPT code (**Figure 6C,D,F**). Finally, we included a few examples of the attention mechanism used in the BERT approach, which highlights some of the many semantic/syntactic dependencies that the model is finding within text subsections (**Figure 7**). These attention matrices were plotted along with importance assigned to subsections of pathology reports using the *All-Fields* model (**Figure 8**), all with their respective textual content.

**Figure 5:**
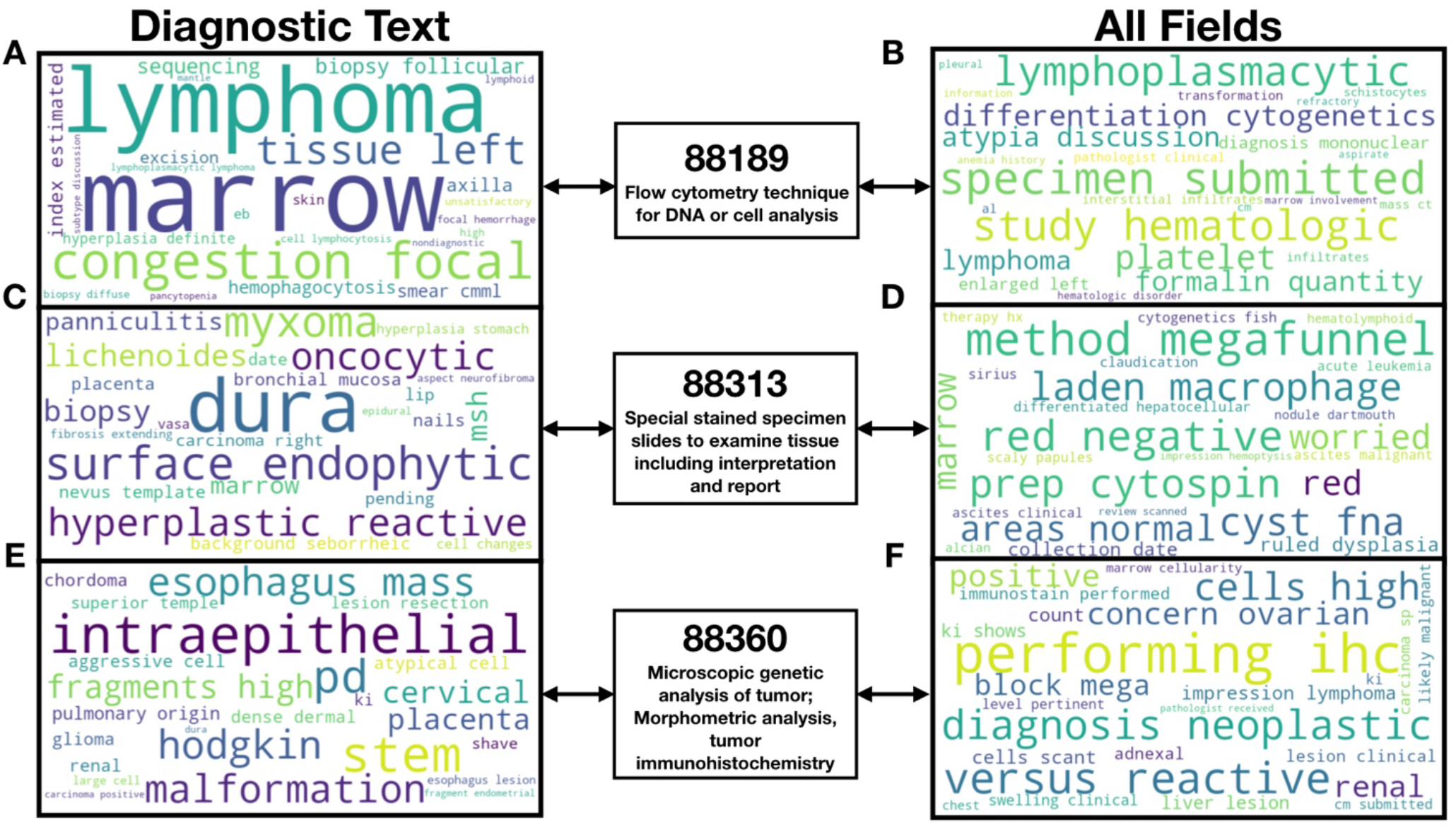
SHAP interpretation of XGBoost predictions: Word clouds demonstrating words found to be important using the XGBoost algorithm for prediction of specific CPT codes, found via shapley attribution; important words pertinent to each CPT code indicated by relative size of the word in the word cloud; word clouds visualized for three example CPT codes: **A-B)** CPT code 88189; **C-D)** CPT code 88313; **E-F)** CPT code 88360; visualizations performed for **A,C,E)** diagnostic text only, **B,D,F)** all report subfields (*all-fields*)

**Figure 6:**
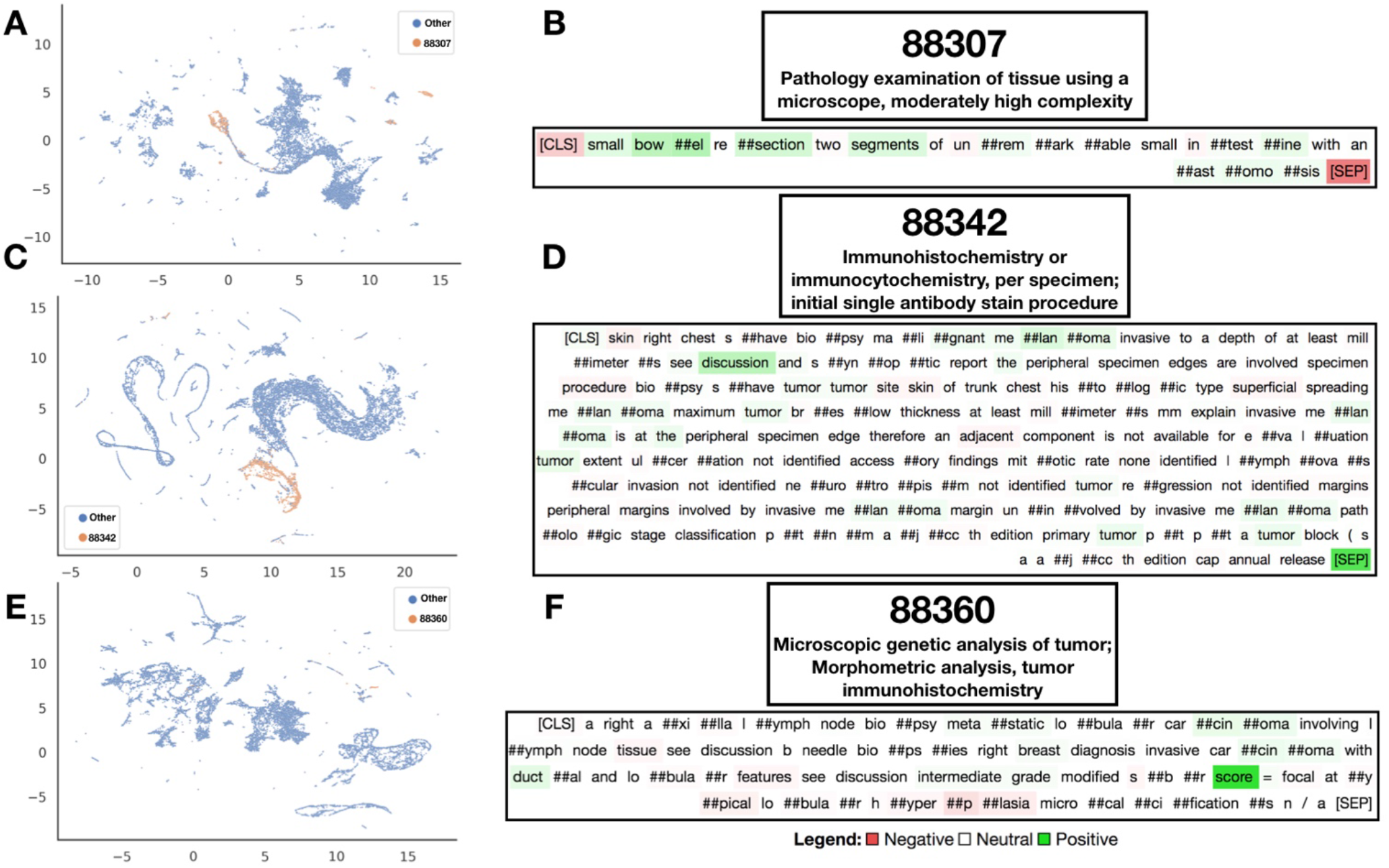
Embedding and Interpretation of BERT Predictions: **A,C,E)** UMAP projection of *All-Fields* BERT embedding vectors after applying attention mechanism across report subfields; each point is report with information aggregated from all report subfields; **B,D,F)** Select diagnostic text from individual reports interpreted by Integrated Gradients to elucidate words positively and negatively associated with calling the CPT code; Integrated Gradients was performed on the diagnostic text BERT models; Utilized CPT codes: **A-B)** CPT code 88307, **C-D)** CPT code 88342, **E-F)** CPT code 88360

**Figure 7:**
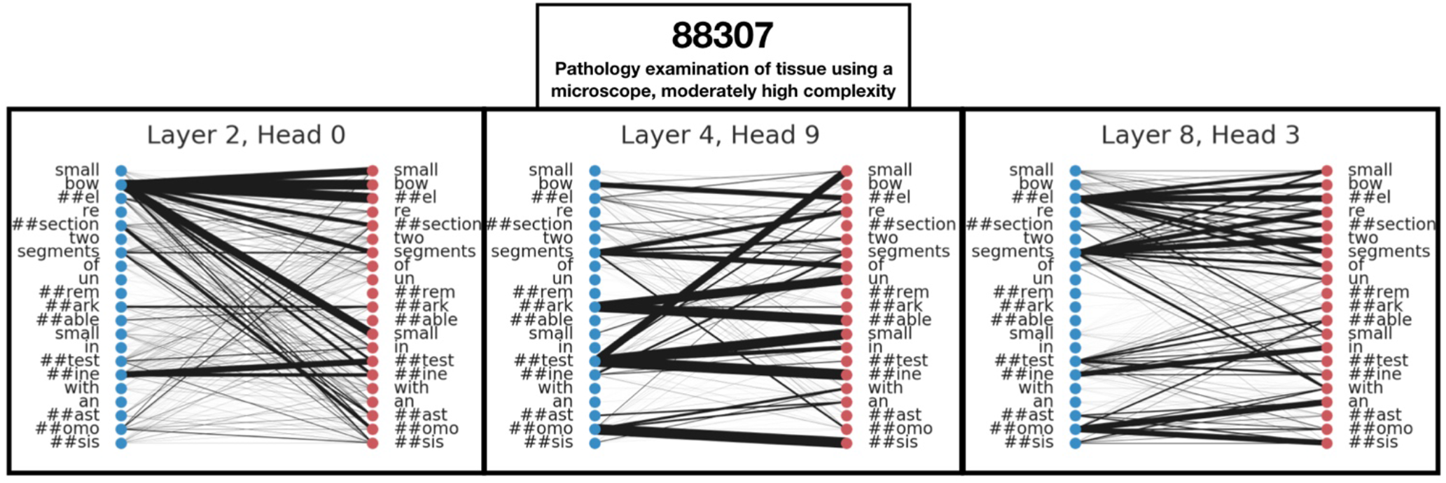
BERT Diagnostic Model Self-Attention: Output of self-attention maps for select self-attention heads/layers from the BERT diagnostic text model visualizes various layers of complex word-to-word relationships for assessment of a select pathology report that was found to report CPT code 88307

**Figure 8:**
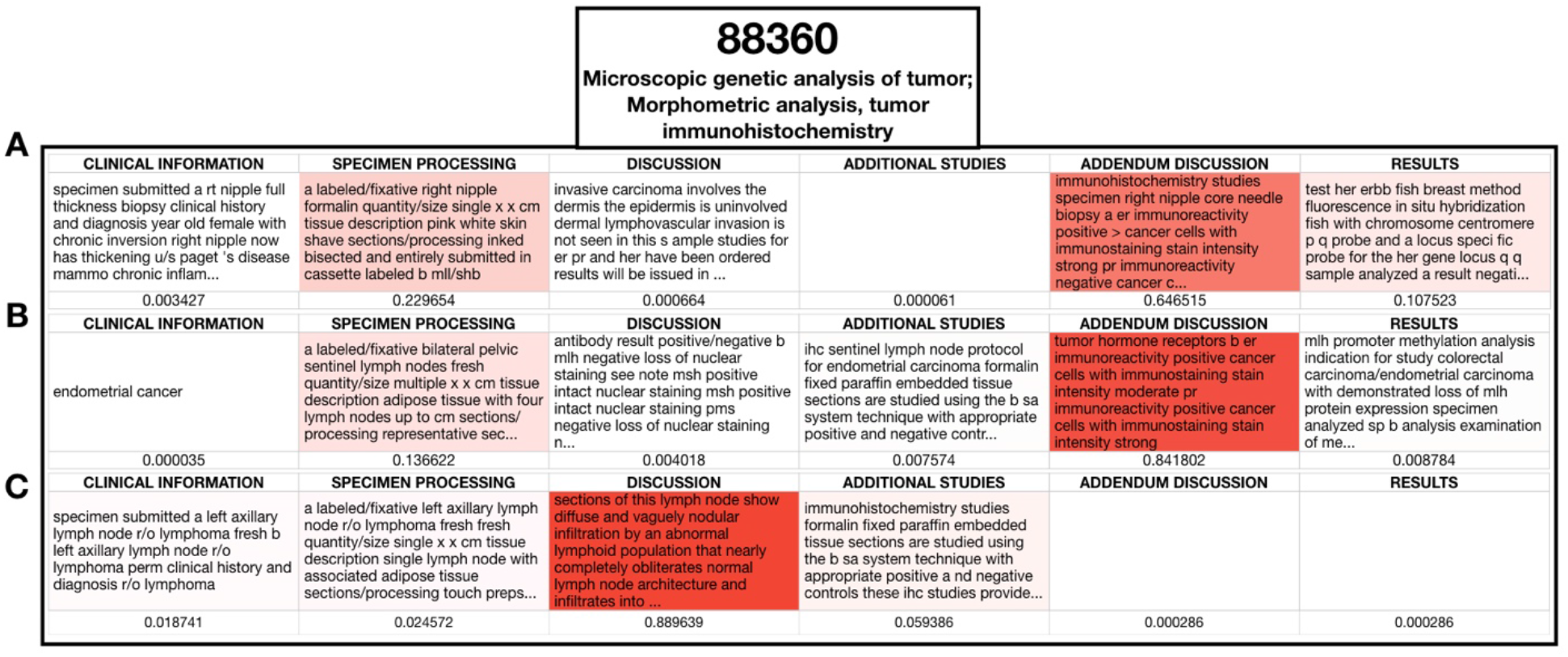
BERT All-Fields Model Interpretation: Visualization of importance scores assigned to pathology report subfields outside of the diagnostic section for three separate pathology reports (**A-C**) that were assigned by raters CPT code 88360; information from report subfields that appear more red were utilized more by the model for the final prediction of the code; attention scores listed below the text from the subfields and title of each subfield supplied

### Pathologist Specific Language

After subsetting to 64,583 documents that are correspondent to the twenty pathologists with the most sign-outs, prediction of the pathologist who had written each pathology report was done with reasonably high accuracy for the XGBoost and BERT approaches. BERT (macro-f1=0.72) performed comparably to XGBoost (macro-f1=0.71) for prediction of pathologist on the diagnostic text; BERT (macro-f1=0.77) and XGBoost (macro-f1=0.78) also performed comparably when considering all report subfields (*all-fields*) (**Supplementary Figure 9**). Model performance improved when incorporating all report subsections. Interestingly, these pathologist-specific subtleties could not be distinguished via the SVM approach (**Supplementary Table 4,8**). Comparisons between the embeddings formed by the *All-Fields* model and that using UMAP (**Figure 9A-B**) shows how the BERT methodology is able to extract features that are more pathologist specific as compared to utilizing a Bag-of-Words approach. Comparing which pathologists were misclassified via the confusion matrix (**Supplementary Figure 6B**) and corroboration with cross tabulations with procedural codes (**Supplementary Figure 6A**) demonstrates that pathologists with similar subspecialties were less distinguishable, however individual patterns persist. We visualized some of the patterns that BERT was able to find in sample sentences via Integrated Gradients and important words via the XGBoost for select pathologists using SHAP (**Figure 10**).

**Figure 9:**
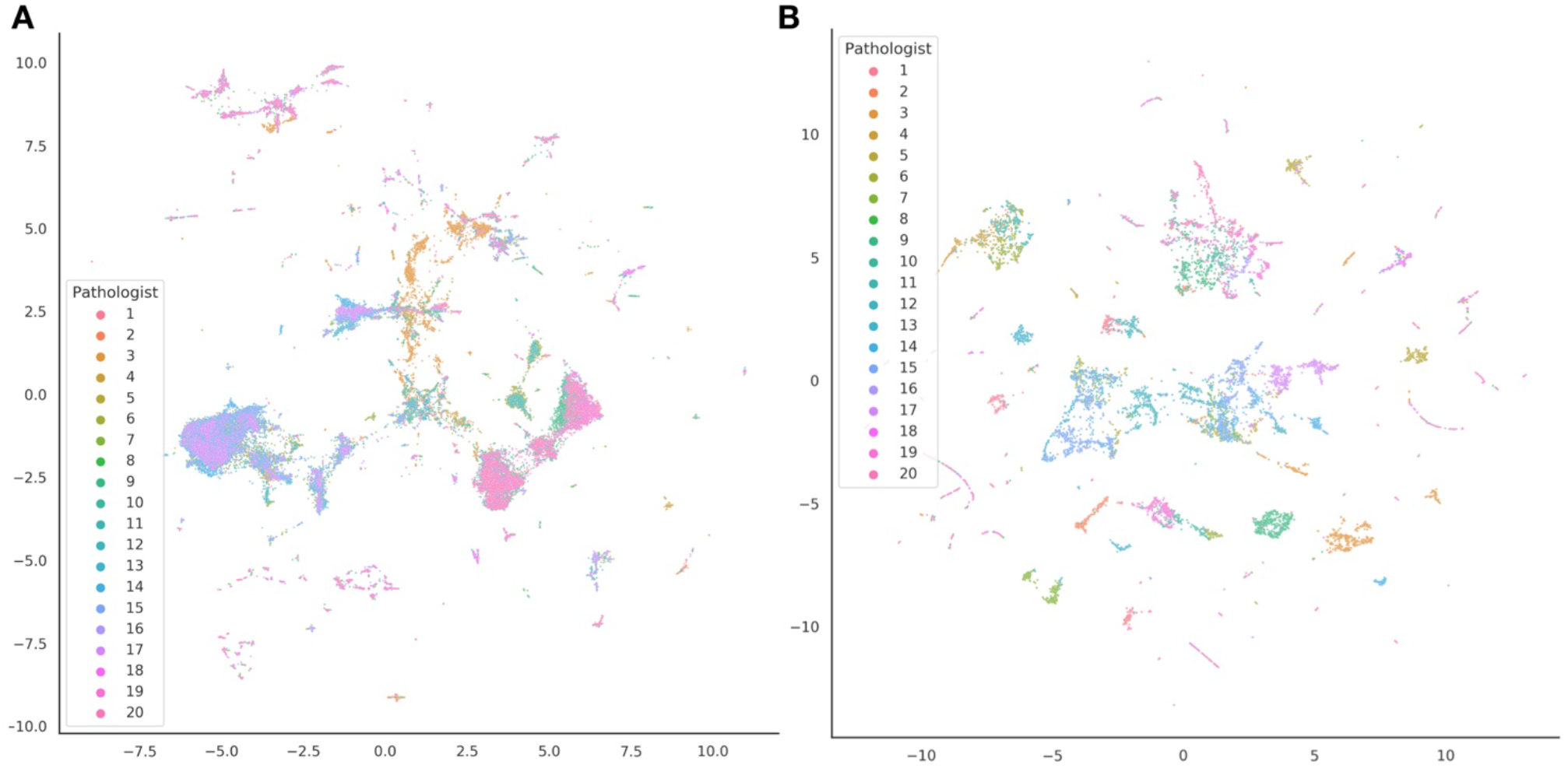
Pathology Reports Colored by Practicing Pathologist: UMAP embeddings of pathology reports, colored by the pathologist who had written the report; each point indicates a pathology report, projected from use of either: **A)** Bag-Of-Words / tf-idf count matrix; **B)** embeddings after integrating information from all report subsections via the BERT *all-fields* model

**Figure 10:**
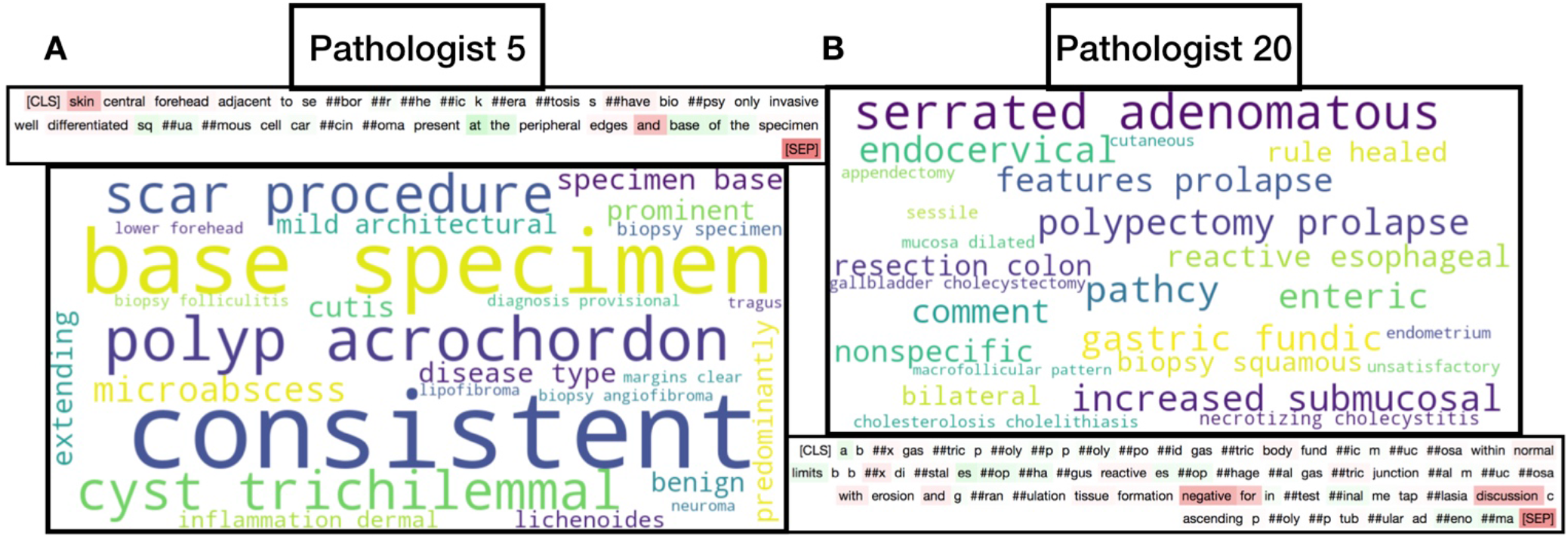
Interpretation of BERT and XGBoost Models for Pathologist Prediction: Word cloud output of top words (size of word indicates importance; importance determined using SHAP) for XGBoost model prediction of specific pathologist and Integrated Gradients highlighting of text via the BERT diagnostic model for select pathologists: **A)** Pathologist 5; **B)** Pathologist 20

## Discussion

In this study, we characterized a large corpus of almost 100,000 pathology reports at a mid-sized academic medical center. We demonstrated that the pathology report subfields contained pertinent diagnostic and procedural information that could adequately separate our text corpus based on CPT code and signing pathologist.. Our studies indicate that the XGBoost and BERT methodologies produce highly accurate predictions of CPT codes, which has the potential to save operating costs by first suggesting codes prior to manual inspection and flagging potential manual coding errors for review. Previous studies predicting CPT codes have largely been unable to characterize the importance of different subsections of a pathology report. Using the BERT method, we were also able to show that significant diagnostic / coding information is contained in non-diagnostic subsections of the pathology report, particularly the Clinical Information and Specimen Processing sections. This is expected, as many of the CPT codes are based on procedure type / specimen complexity. Furthermore, we were able to assess nuanced pathologist specific language, which was largely determined by specialty (e.g. subspecialities like cytology use highly regimented language making it more difficult to separate practitioners). While our prediction accuracy is comparable to previous reports of CPT prediction using machine learning methods, our work covers a wider range of codes than previously reported, compares the different algorithms through rigorous cross-validation, reports significantly higher sensitivity and specificity, and demonstrates the importance of utilizing other parts of the pathology report for procedural code prediction.

It was interesting to note how some of the clinical codes for acquisition and quantification of markers on specialized stains (CPT 88341, 88342, 88344, 88360) performed the worst overall. The revision of CPT codes 88342 and 88360, and addition of CPT codes 88341 and 88344 in 2015 lay just outside of the range of the data collection period, which was from June 2015 to June 2020 ^50^. Evolving coding/billing guidelines will always present challenges when developing NLP guidelines for clinical tests, though results did indicate that overall, temporal changes in coding did not significantly affect the ability to predict codes using a wide variety of machine learning techniques. Since major improvements were obtained through incorporating the other report subfields for the codes, non-diagnostic text may be more important for records of specialized stain processing and should be utilized as such.

## Limitations

There are a few limitations to our work. For instance, due to computational constraints, most BERT models can only take as input 512 words at a time. We utilized a pretrained BERT model that inherited knowledge from large existing biomedical data repositories at the expense of flexibility in sequence length size (i.e. we could not modify the word limit while utilizing this pre-trained model). We noticed that in our text corpus, less than 2% of reports were longer than this limitation and thus had to be truncated when input into the deep learning model, which may impact results. Potentially, longer pathology reports describe more complicated cases, of which may utilize additional procedures. From our cluster analysis, we demonstrated that this appeared to be the case for a subset of report clusters, though for one cluster, the opposite was true. However, a vast majority of pathology reports fell within the BERT word limits, so we considered any word length based association with CPT code complexity to have negligible impact on the model results. The XGBoost model, alternatively, is able to operate on the entire report text. Thus, XGBoost may more directly capture interactions between words spanning across document subsections pertaining to complex cases, which may serve as one plausible explanation of its apparent performance increase with respect to the deep learning approaches. Due to the significant compute time on many of these algorithms, we performed limited hyperparameter tuning.

### Future Directions

While much of the patient’s narrative may be told separately through text, imaging, and omics modalities ^51^, there is tremendous potential to integrate semantic information contained in pathologist notes with imaging and omics modalities to capture a more holistic perspective of the patient’s health and integrate potentially useful information that could otherwise be overlooked. For instance, the semantic information contained in a report may highlight specific morphological and macro-architectural features in the correspondent biopsy specimen that an image-based deep learning model might struggle to identify without additional information.

Although XGBoost demonstrated equivalent performance with the deep learning methods used for CPT prediction, its usefulness in a multimodal model is limited because these machine learning approaches rely heavily on the feature extraction approach, where feature generation mechanisms using deep learning can be tweaked during optimization to complement the other modalities. Alternatively, the semantic information contained within the word embedding layers of the BERT model can be fine-tuned when used in conjunction with or directly predicting on imaging data allowing for more seamless integration of multi-omic data. There is also potentially useful information to be gained by working to identify text that can distinguish pathologists within subspecialties rather than identify pathologists across subspecialties. This information can be useful in helping to create more standardized lexicons / diagnostic rubrics (for instance, The Paris System for Urine Cytopathology ^52^). Research into creating a standard lexicon for particular specialties or converting raw free text into a standardized report could be very fruitful especially for the positive impact it would have in allowing non-pathologist physicians to more easily interpret pathology reports and make clinical decisions.

We are also interested in investigations of outlier text as a marker of uncertainty. For instance, if there is a text content outlier in a body of reports with the same CPT code then we can hypothesize that such text may be more prone to ambiguous phrases or hedging, from which pathologists may articulate their uncertainty for a definitive diagnosis. We would also like to assess the impact of hedging in the assignment of procedural codes, and furthermore its subsequent impact on patient care. To ameliorate these differences in reporting patterns, generative deep learning methods can be employed to summarize the text through generation of a standard lexicon. Other excellent applications of BERT-based text models include prediction of relative value units (RVU’s) via report complexity for pathologist compensation calculations and detection of cases that may have been mis-billed which can potentially save the hospital resources. We also plan to incorporate newer deep learning architectures, such as the Reformer or Albert, which do not suffer from the word length limitations of BERT, though training all possible language models was outside of the scope of our study.

## Conclusion

In this study, we compare three cutting-edge machine learning techniques for the prediction of CPT codes from pathology text. Our results provide additional evidence for the utility of machine learning models to predict CPT codes in a large corpus of pathology reports acquired from a mid-sized academic medical center. Furthermore, we demonstrated that utilizing text from parts of the document other than the diagnostic section aids in the extraction of procedural information. While the XGBoost and BERT methodologies both yielded comparable results, either method can be used to improve the speed and accuracy of coding by suggestion of relevant CPT codes to coders, though deep learning approaches present the most viable methodology for incorporating text data with other pathology modalities.

## Data Availability

The EHR dataset curated from Dartmouth-Hitchcock records contains information that
could compromise research participant privacy/consent and thus cannot be released
due to HIPAA regulations. An IRB approval is required for on-site access and review of
the data.

## Notes

### Competing Interest Statement

The authors have declared no competing interest.

### Funding Statement

This work was supported by:
NIH grants R01CA216265, R01DE022772, and P20GM104416 to BCC.
Two Dartmouth College Neukom Institute for Computational Science CompX awards to BCC, LJV and JJL.
JJL is supported through the Burroughs Wellcome Fund Big Data in the Life Sciences training grant at Dartmouth.
Norris Cotton Cancer Center, DPLM Clinical Genomics and Advanced
Technologies EDIT program.
The funding bodies above did not have any role in the study design, data collection, analysis and interpretation, or writing of the manuscript.

### Author Declarations

We obtained an Institutional Review Board approval from Dartmouth Hitchcock Medical Center.

