## Supplementary Materials for "Comparison of Machine Learning Algorithms for the Prediction of Current Procedural Terminology (CPT) Codes from Pathology Reports"

**Supplementary Table 1:** Recording the percent missingness of each report subsection before removing reports lacking a diagnostic section. Summary measures (median, 1st quartile, 3rd quartile) for number of words in each document subsection (where the subfield existed) and the percentage of documents whose length exceeded 512 words.

| Report Subfield | Missingness Before Removal | Median Word Count | 1st Q Word Count | 3rd Q Word Count | Exceeds BERT Max Words |
|---|---|---|---|---|---|
| **ADDENDUM DISCUSSION** | 96.2% | 84 | 47 | 121 | 0.000% |
| **ADDITIONAL STUDIES** | 86.6% | 78 | 11 | 92 | 0.039% |
| **CLINICAL INFORMATION** | 5.3% | 25 | 18 | 61 | 0.000% |
| **DIAGNOSIS** | 3.5% | 23 | 13 | 28 | 0.022% |
| **DISCUSSION** | 81.7% | 36 | 16 | 68 | 0.017% |
| **FINAL DIAGNOSIS** | 99.9% | 235 | 186 | 313 | 0.000% |
| **FROZEN SECTION** | 99.4% | 2 | 1 | 11 | 0.000% |
| **FROZEN SECTION DIAGNOSIS** | 99.3% | 20 | 12 | 33 | 0.000% |
| **INTERPRETATION** | 99.9% | 135 | 91 | 141 | 0.000% |
| **RESULTS** | 97.9% | 248 | 216 | 268 | 2.198% |
| **SPECIMEN PROCESSING** | 34.4% | 38 | 27 | 64 | 0.389% |
| **Complete Text (*All Fields*)** | 0% | 119 | 68 | 158 | 1.768% |

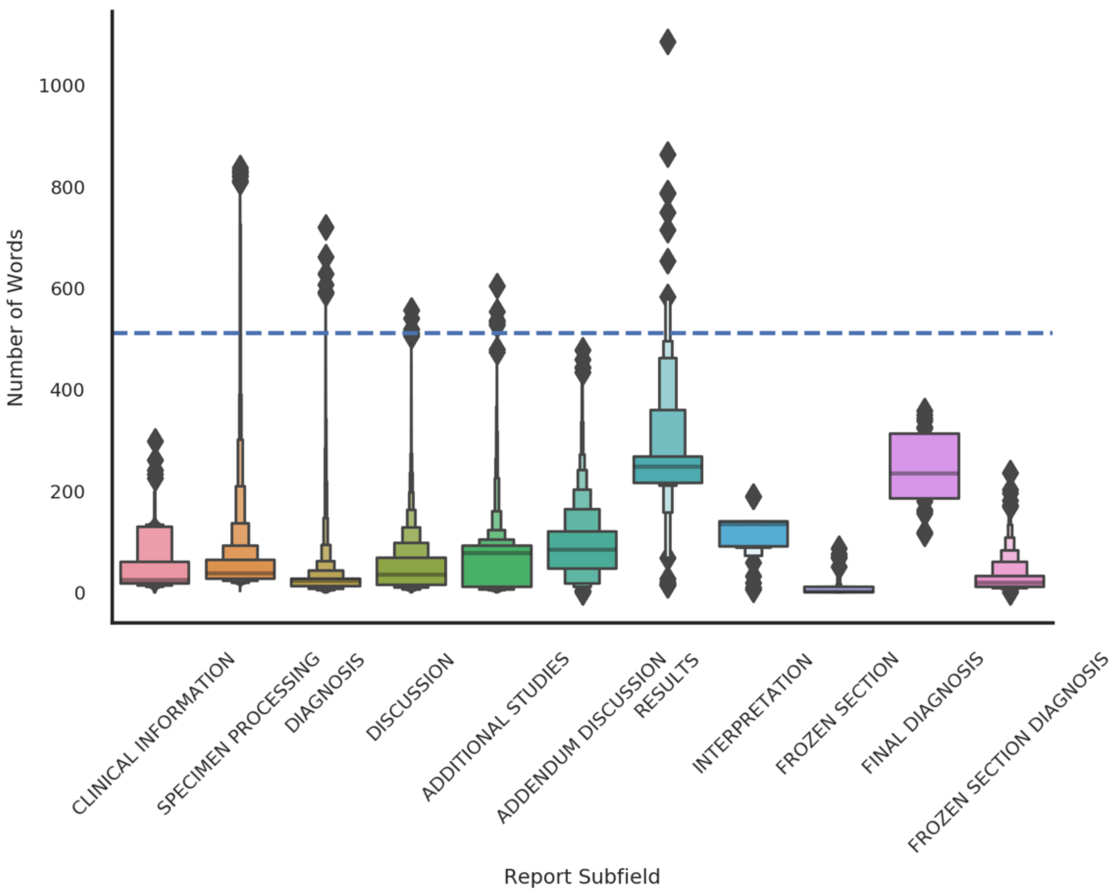

**Supplementary Figure 1:** Boxenplots of number of words for each subfield across pathology report corpus; BERT cutoff word count of 512 words represented by horizontal dashed line

**Supplementary Table 2:** Correlation between length of the word document and the number of uniquely assigned codes; broken down by reported cluster using the diagnostic fields and all report fields

| | ***Diagnostic Clusters*** | | ***All-Fields Clusters*** | |
|---|---|---|---|---|
| ***Cluster*** | **Correlation** | **p-value** | **Correlation** | **p-value** |
| ***1*** | -0.09 | 1.6E-26 | 0.39 | 5.7E-178 |
| ***2*** | 0.07 | 1.6E-02 | -0.05 | 7.1E-02 |
| ***3*** | -0.30 | 3.4E-85 | 0.01 | 7.1E-01 |
| ***4*** | -0.05 | 2.8E-02 | 0.02 | 3.9E-01 |
| ***5*** | 0.18 | 6.9E-93 | 0.00 | 9.6E-01 |
| ***6*** | 0.21 | 9.4E-37 | 0.01 | 2.9E-01 |
| ***7*** | 0.27 | 2.5E-26 | 0.08 | 1.7E-05 |
| ***8*** | 0.14 | 6.3E-137 | 0.57 | 4.9E-93 |
| ***9*** | | | 0.10 | 1.0E-28 |
| ***10*** | | | 0.31 | 5.3E-113 |
| ***11*** | | | 0.32 | 1.2E-33 |
| ***12*** | | | 0.16 | 1.0E-23 |
| ***13*** | | | 0.48 | 5.1E-98 |
| ***14*** | | | 0.09 | 3.9E-07 |
| ***15*** | | | 0.32 | 0.0E+00 |

**Supplementary Table 3:** Top ten words found for each LDA topic ("topic descriptors"); Ten topics were discovered for the diagnostic text; Ten additional topics were discovered for all of the report subfields (*All Fields*)

| Diagnostic Text | Topic 1 | Topic 2 | Topic 3 | Topic 4 | Topic 5 | Topic 6 | Topic 7 | Topic 8 | Topic 9 | Topic 10 |
|---|---|---|---|---|---|---|---|---|---|---|
| **0** | tumor | tissue | test | mucosa | cervical | colon | nevus | cells | shave | fragments |
| **1** | lymph | right | cancer | gastric | results | polypectomy | shave | placenta | cell | benign |
| **2** | carcinoma | left | lesion | esophagus | cancer | tubular | excision | cord | carcinoma | endocervical |
| **3** | grade | benign | cervical | chronic | please | adenoma | left | umbilical | left | evidence |
| **4** | nodes | excision | please | normal | guidelines | polyp | right | vessel | right | cervical |
| **5** | prostatic | soft | results | within | test | hyperplastic | melanocytic | acute | specimen | effect |
| **6** | left | breast | consensus | limits | consensus | ascending | changes | seen | basal | squamous |
| **7** | right | fallopian | management | abnormality | screening | sigmoid | compound | three | squamous | dysplasia |
| **8** | identified | inflammation | http://www.asccp.org | diagnostic | management | fragments | specimen | grams | discussion | mucosa |
| **9** | invasive | resection | guidelines | seen | cells | transverse | back | villous | peripheral | hpv |
| **All Fields Text** | **Topic 1** | **Topic 2** | **Topic 3** | **Topic 4** | **Topic 5** | **Topic 6** | **Topic 7** | **Topic 8** | **Topic 9** | **Topic 10** |
| **0** | pap | skin | tissue | tissue | pap | tissue | biopsy | positive | clinical | skin |
| **1** | hpv | specimen | biopsy | polyp | hist | lymph | diagnosis | antibody | pertinent | shave |
| **2** | test | clinical | formalin | submitted | hpv | margin | specimen | tissue | total | biopsy |
| **3** | hist | 'clock | quantity/size | colon | test | specimen | clinical | clinical | received | left |
| **4** | screening | excision | sections/processing | formalin | screening | tumor | see | studies | fluid | clinical |
| **5** | cervical | submitted | submitted | clinical | cervical | right | case | diagnostic | source | right |
| **6** | clinical | tissue | labeled/fixative | soft | clinical | left | punch | formalin | specimen | submitted |
| **7** | therapy | left | description | labeled/fixative | therapy | node | discussion | staining | preparation | specimen |
| **8** | cancer | nevus | soft | history | cancer | submitted | submitted | core | description | tissue |

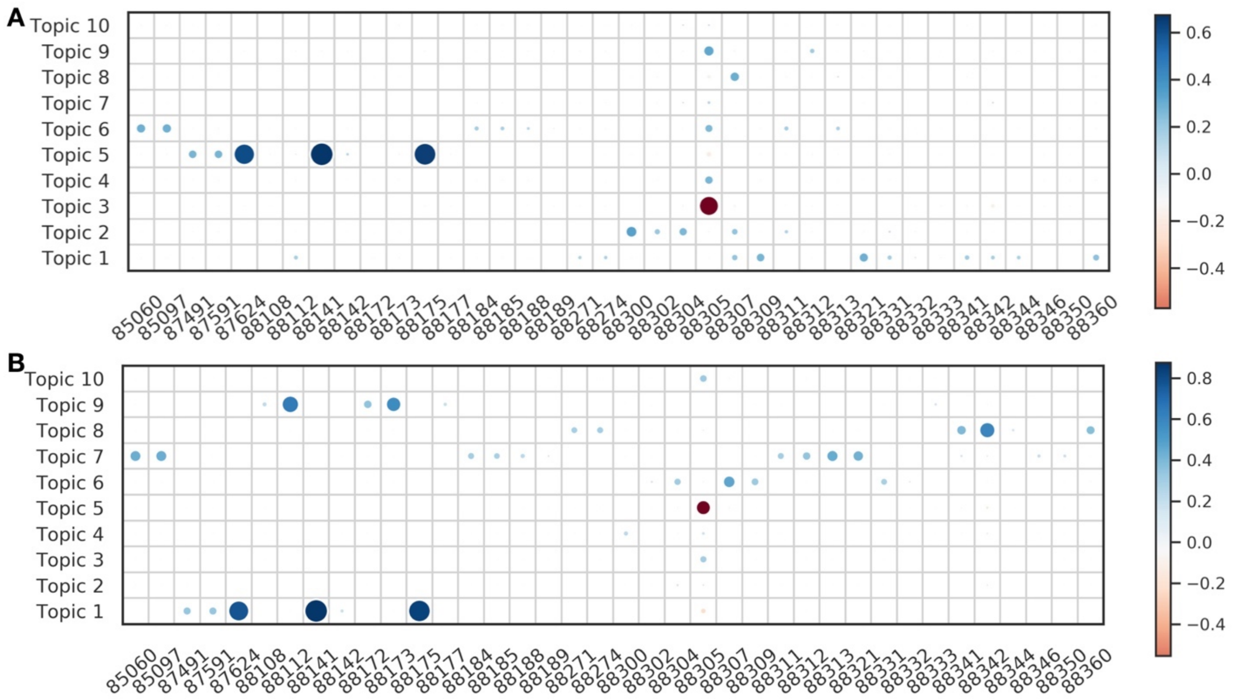

**Supplementary Figure 2:** Strength of correlation between topics and CPT codes denoted by size and color of each circle; large blue circles indicate strong positive associations, while large red circles indicate strong negative associations; associations for: **A)** diagnostic text; **B)** all-fields text

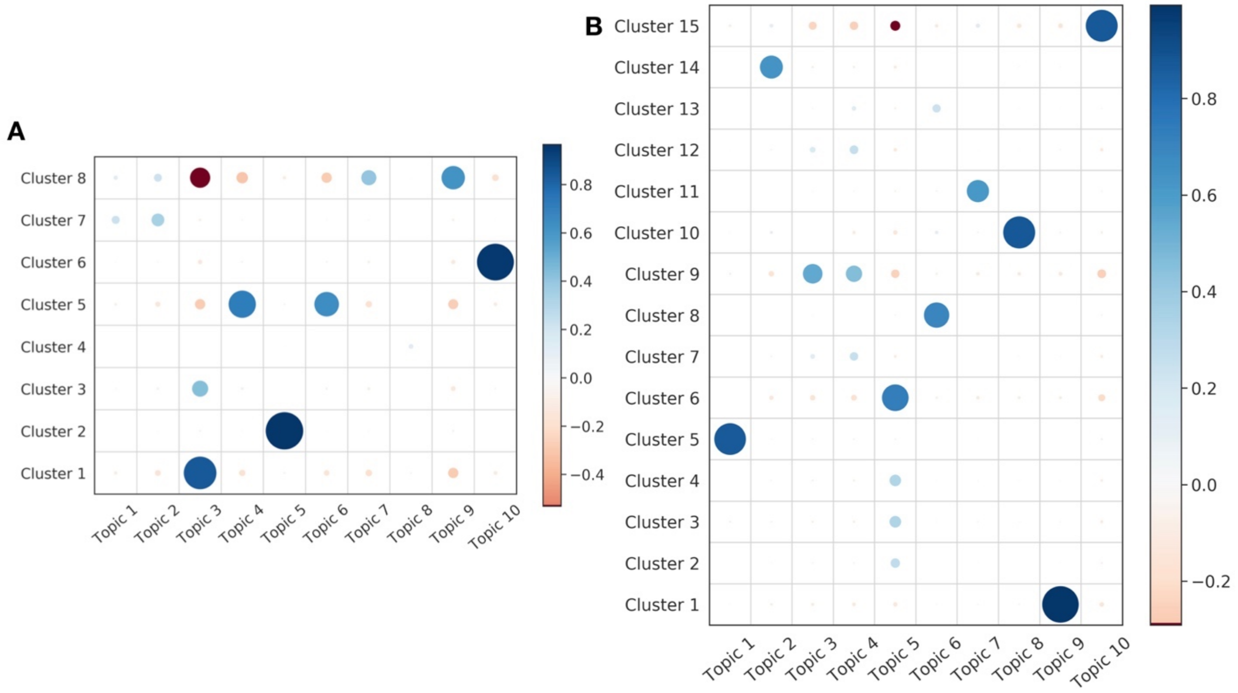

**Supplementary Figure 3:** Strength of correlation between topics and HDBSCAN report clusters denoted by size and color of each circle; large blue circles indicate strong positive associations, while large red circles indicate strong negative associations; associations for: **A)** diagnostic text; **B)** all-fields text

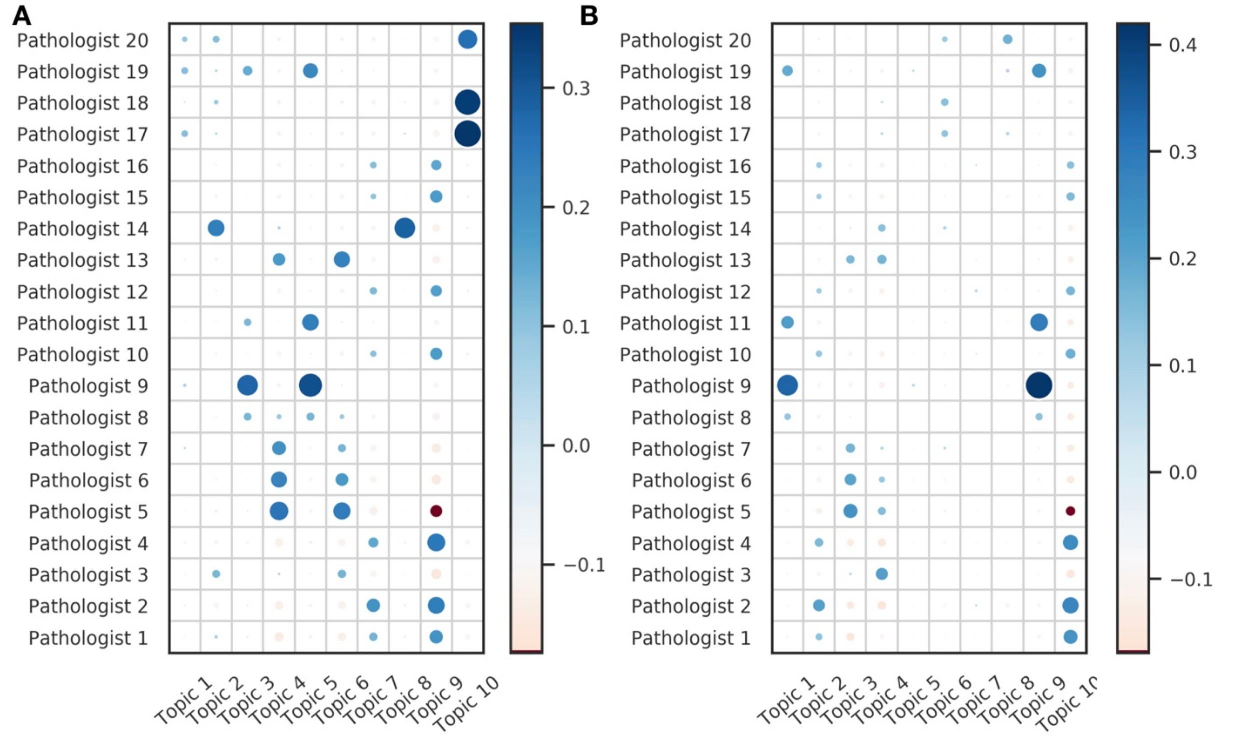

**Supplementary Figure 4:** Strength of correlation between topics and individual pathologists denoted by size and color of each circle; large blue circles indicate strong positive associations, while large red circles indicate strong negative associations; associations for: **A)** diagnostic text; **B)** all-fields text

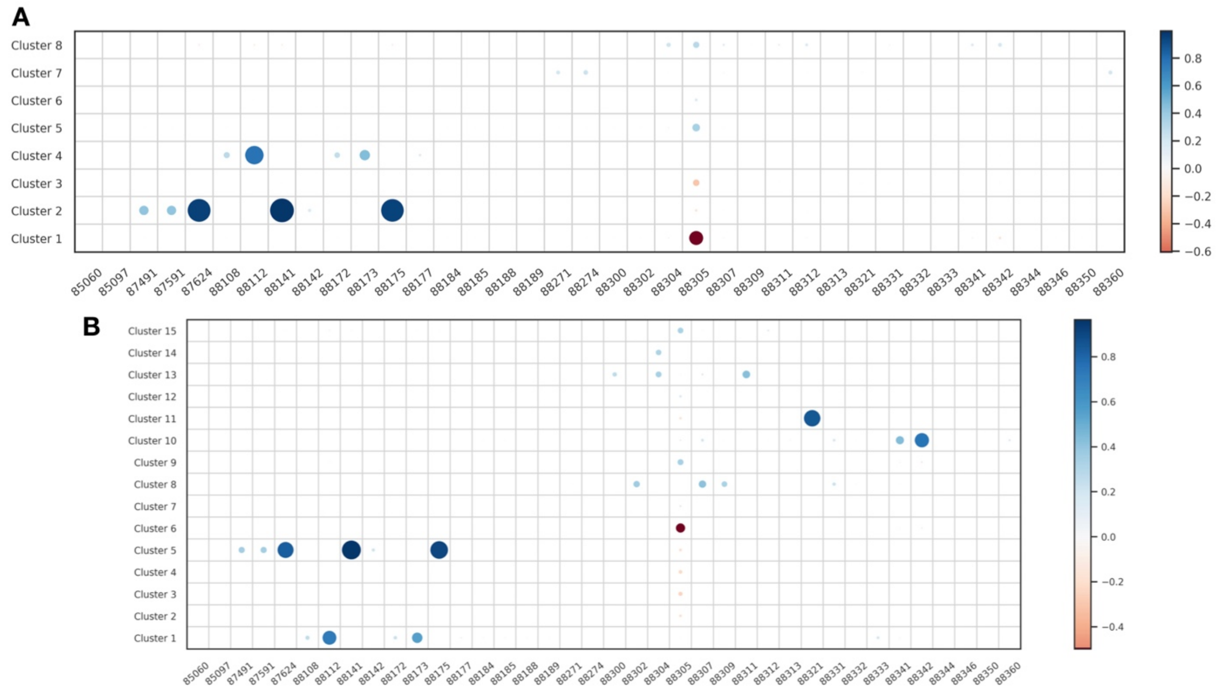

**Supplementary Figure 5:** Strength of correlation between CPT codes and HDBSCAN report clusters denoted by size and color of each circle; large blue circles indicate strong positive

associations, while large red circles indicate strong negative associations; associations for: **A)** diagnostic text; **B)** all-fields text

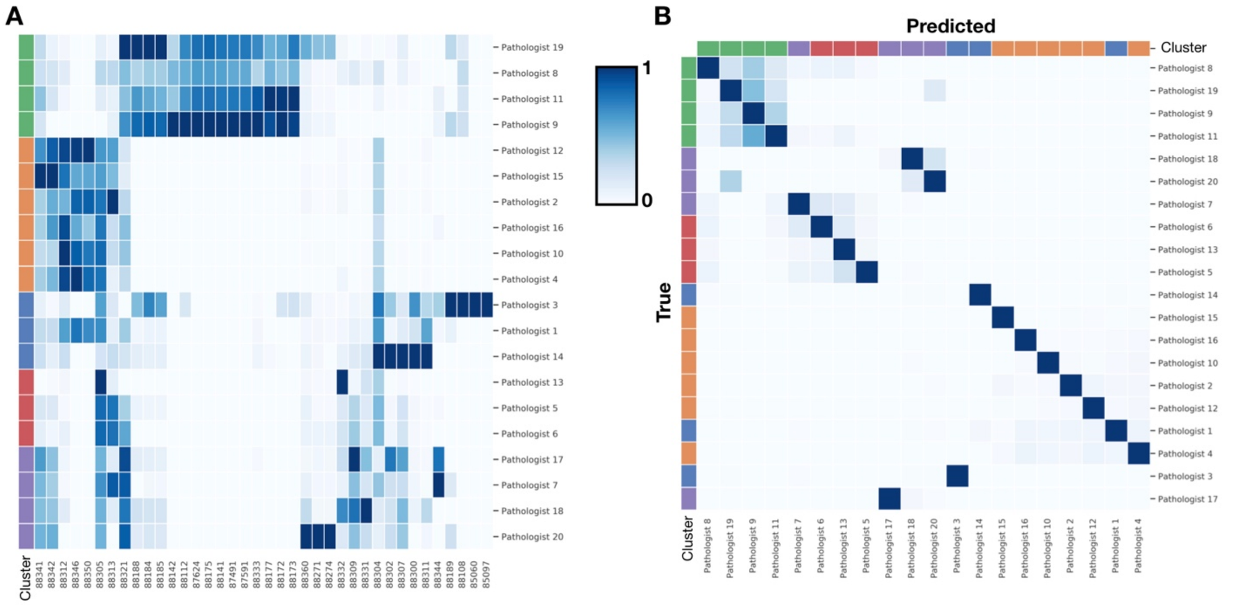

**Supplementary Figure 6:** Pathologist Associations: **A)** Clustered heatmap between associations/co-occurence between pathologist and CPT codes establishes “subspecialties”, where pathologists who order similar CPT codes are likely of similar subspecialty/subspecialties; left color track is colored by established subspecialty clusters; **B)** Clustered confusion matrix for pathologist prediction task (BERT diagnostic-text model); rows indicate true pathologists, while columns indicate predicted pathologist; row and column color bars utilize established “subspecialty” clusters; since clustering of rows and columns place pathologists of similar subspecialty together, this indicates that misclassification occurred mostly within subspecialty

**Supplementary Table 4:** Confidence intervals of 1000-sample non-parametric bootstrap of area under the receiver operating characteristic curve for each algorithm (BERT, XGBoost and SVM) and for each report type (Diagnosis and *All-Fields*); each AUC was averaged across the 5-cross validation folds with same random seed set for sampling values within each CV fold for each code/group of pathologists; CPT code and descriptions of codes listed on left in addition to weighted AUC across 20 pathologists

| | | **AUCs (±SE)** | | | | | |
|---|---|---|---|---|---|---|---|
| | | **BERT** | | **XGBoost** | | **SVM** | |
| **Code** | **Description** | **Diagnosis** | **All-Fields** | **Diagnosis** | **All-Fields** | **Diagnosis** | **All-Fields** |
| **85060** | **Blood smear interpretation by physician with written report** | 0.998±0.0002 | 0.9994±0.0001 | 0.9989±0.0002 | 0.9996±0.0001 | 0.9983±0.0002 | 0.9968±0.0012 |
| **85097** | **Bone marrow, smear interpretation** | 0.9996±0.0001 | 0.9994±0.0001 | 0.9989±0.0005 | 0.9997±0.0 | 0.9985±0.0001 | 0.9941±0.0014 |
| **87491** | **Detection test for chlamydia** | 0.9905±0.0008 | 0.9984±0.0008 | 0.9898±0.001 | 0.9996±0.0002 | 0.9872±0.0013 | 0.9819±0.0042 |
| **87591** | **Detection test for Neisseria gonorrhoeae (gonorrhoeae bacteria)** | 0.9905±0.0008 | 0.9994±0.0001 | 0.9898±0.001 | 0.9996±0.0002 | 0.9872±0.0013 | 0.9819±0.0042 |

| | | | | | | | |
|---|---|---|---|---|---|---|---|
| **87624** | **Detection test for human papillomavirus (hpv)** | 0.9968±0.0006 | 0.9973±0.0003 | 0.9958±0.0004 | 0.9984±0.0002 | 0.9778±0.0017 | 0.988±0.0016 |
| **88108** | **Cell examination of specimen** | 0.9802±0.0017 | 0.999±0.0003 | 0.9808±0.0008 | 0.9975±0.0015 | 0.9717±0.0026 | 0.9989±0.0001 |
| **88112** | **Cell examination of specimen** | 0.9934±0.0005 | 0.9991±0.0001 | 0.9935±0.0002 | 0.9995±0.0 | 0.9887±0.0004 | 0.9959±0.0008 |
| **88141** | **Cytopathology, cervical or vaginal (any reporting system), requiring interpretation by physician** | 1.0±0.0 | 0.9998±0.0001 | 0.9996±0.0001 | 0.9999±0.0 | 0.9988±0.0004 | 0.9923±0.0014 |
| **88142** | **Pap test (Pap smear)** | 0.9886±0.0017 | 0.9938±0.0016 | 0.9826±0.0017 | 0.9951±0.0018 | 0.9663±0.0018 | 0.9501±0.0131 |
| **88172** | **Evaluation of fine needle aspirate** | 0.9825±0.0011 | 0.999±0.0002 | 0.9837±0.0011 | 0.999±0.0006 | 0.9749±0.0015 | 0.9903±0.001 |
| **88173** | **Evaluation of fine needle aspirate with interpretation and report** | 0.9867±0.0024 | 0.9988±0.0002 | 0.9899±0.0005 | 0.9996±0.0 | 0.9818±0.001 | 0.997±0.0006 |
| **88175** | **Pap test** | 0.998±0.0005 | 0.9976±0.0003 | 0.9972±0.0003 | 0.9981±0.0003 | 0.9932±0.0009 | 0.9847±0.002 |
| **88177** | **Pap Test** | 0.9774±0.0023 | 0.9993±0.0001 | 0.9783±0.0031 | 0.9998±0.0 | 0.9624±0.0044 | 0.9955±0.0003 |
| **88184** | **Flow cytometry technique for DNA or cell analysis** | 0.9731±0.0082 | 0.9848±0.0022 | 0.9738±0.0025 | 0.9942±0.0012 | 0.9699±0.0029 | 0.9708±0.0033 |
| **88185** | **Flow cytometry, cell suXGBace, cytoplasmic, or nuclear marker, technical component only** | 0.9629±0.0075 | 0.9841±0.0022 | 0.9711±0.0027 | 0.994±0.0008 | 0.9594±0.003 | 0.9692±0.0034 |
| **88188** | **Cytopathology Procedures.** | 0.9428±0.0121 | 0.9773±0.0029 | 0.9589±0.0041 | 0.9875±0.0024 | 0.9593±0.0041 | 0.9486±0.0029 |
| **88189** | **Flow cytometry technique for DNA or cell analysis** | 0.9043±0.0295 | 0.9753±0.0052 | 0.9199±0.0101 | 0.9785±0.0073 | 0.9611±0.0074 | 0.9471±0.0118 |
| **88271** | **FISH DNA probe, each** | 0.9943±0.002 | 0.9906±0.0025 | 0.9735±0.0055 | 0.995±0.0024 | 0.9717±0.0062 | 0.9768±0.0061 |
| **88274** | **Genetic testing** | 0.9951±0.0011 | 0.9943±0.003 | 0.9755±0.0059 | 0.9941±0.0036 | 0.9775±0.0058 | 0.9922±0.0029 |
| **88300** | **Pathology examination of tissue using a microscope, limited examination** | 0.9983±0.0011 | 0.9969±0.0008 | 0.9967±0.0012 | 0.9978±0.0009 | 0.9846±0.0025 | 0.9868±0.0023 |
| **88302** | **Pathology examination of tissue using a microscope** | 0.9768±0.0083 | 0.9824±0.0036 | 0.9887±0.0028 | 0.9934±0.0019 | 0.9581±0.0047 | 0.9643±0.0042 |
| **88304** | **Pathology examination of tissue using a microscope, moderately low complexity** | 0.991±0.0011 | 0.9877±0.0007 | 0.987±0.0009 | 0.9907±0.0006 | 0.9534±0.0019 | 0.9509±0.0021 |
| **88305** | **Pathology examination of tissue using a microscope, intermediate complexity** | 0.9726±0.0012 | 0.9775±0.0005 | 0.97±0.0006 | 0.9889±0.0003 | 0.1087±0.0012 | 0.0807±0.001 |
| **88307** | **Pathology examination of tissue using a microscope, moderately high complexity** | 0.9942±0.0006 | 0.9928±0.0004 | 0.9925±0.0004 | 0.995±0.0003 | 0.9614±0.0015 | 0.968±0.0013 |
| **88309** | **Pathology examination of tissue using a microscope, high complexity** | 0.9966±0.0009 | 0.9885±0.0021 | 0.9949±0.0008 | 0.9967±0.0007 | 0.9608±0.0034 | 0.9777±0.0022 |

| | | | | | | | |
|---|---|---|---|---|---|---|---|
| **88311** | **Preparation of tissue for examination by removing any calcium present** | 0.9906±0.0033 | 0.9972±0.0003 | 0.9943±0.0009 | 0.9991±0.0002 | 0.9316±0.0035 | 0.9741±0.0019 |
| **88312** | **Special stained specimen slides to identify organisms including interpretation and report** | 0.9766±0.0025 | 0.9792±0.0012 | 0.9692±0.0017 | 0.9972±0.0004 | 0.8974±0.0038 | 0.9063±0.0031 |
| **88313** | **Special stained specimen slides to examine tissue including interpretation and report** | 0.9577±0.0065 | 0.9854±0.0013 | 0.9619±0.0023 | 0.9953±0.0006 | 0.9163±0.0039 | 0.9234±0.0036 |
| **88321** | **Surgical pathology consultation and report** | 0.9945±0.0007 | 0.998±0.0007 | 0.9889±0.001 | 0.9994±0.0001 | 0.9483±0.0033 | 0.9931±0.0013 |
| **88331** | **Pathology examination of tissue during surgery** | 0.949±0.0135 | 0.9958±0.0012 | 0.9834±0.0019 | 0.9996±0.0002 | 0.9465±0.0044 | 0.9592±0.0024 |
| **88332** | **Pathology examination of specimen during surgery** | 0.8971±0.0485 | 0.9821±0.0063 | 0.974±0.0059 | 0.9972±0.0008 | 0.9077±0.0186 | 0.9666±0.0084 |
| **88333** | **Pathology examination of tissue specimen during surgery** | 0.9924±0.0011 | 0.9963±0.0018 | 0.9883±0.0027 | 0.999±0.0008 | 0.9827±0.0021 | 0.979±0.0076 |
| **88341** | **Immunohistochemistry or immunocytochemistry, per specimen** | 0.9353±0.0034 | 0.96±0.0012 | 0.9273±0.0017 | 0.9901±0.0004 | 0.8514±0.0031 | 0.9262±0.0022 |
| **88342** | **Immunohistochemistry or immunocytochemistry, per specimen; initial single antibody stain procedure** | 0.9384±0.0024 | 0.9925±0.0003 | 0.9319±0.0011 | 0.9955±0.0002 | 0.8404±0.0021 | 0.9471±0.0015 |
| **88344** | **Special stained specimen slides to examine tissue** | 0.9833±0.0117 | 0.9824±0.0075 | 0.9747±0.0061 | 0.9942±0.0028 | 0.9664±0.0091 | 0.9627±0.0091 |
| **88346** | **Antibody evaluation** | 0.9971±0.0028 | 0.9972±0.0018 | 0.9966±0.0026 | 0.9977±0.0023 | 0.987±0.0045 | 0.989±0.005 |
| **88350** | **Antibody evaluation** | 0.9999±0.0001 | 0.9998±0.0 | 0.9993±0.0004 | 0.9999±0.0 | 0.9852±0.0048 | 0.9933±0.0037 |
| **88360** | **Microscopic genetic analysis of tumor; Morphometric analysis, tumor immunohistochemistry** | 0.7182±0.0282 | 0.9853±0.0022 | 0.9761±0.0027 | 0.9944±0.0013 | 0.9578±0.0042 | 0.9564±0.0048 |
| | **Top 20 Pathologists** | 0.984±0.0002 | 0.9877±0.0002 | 0.9823±0.0002 | 0.99±0.0001 | 0.3778±0.0007 | 0.3726±0.0007 |

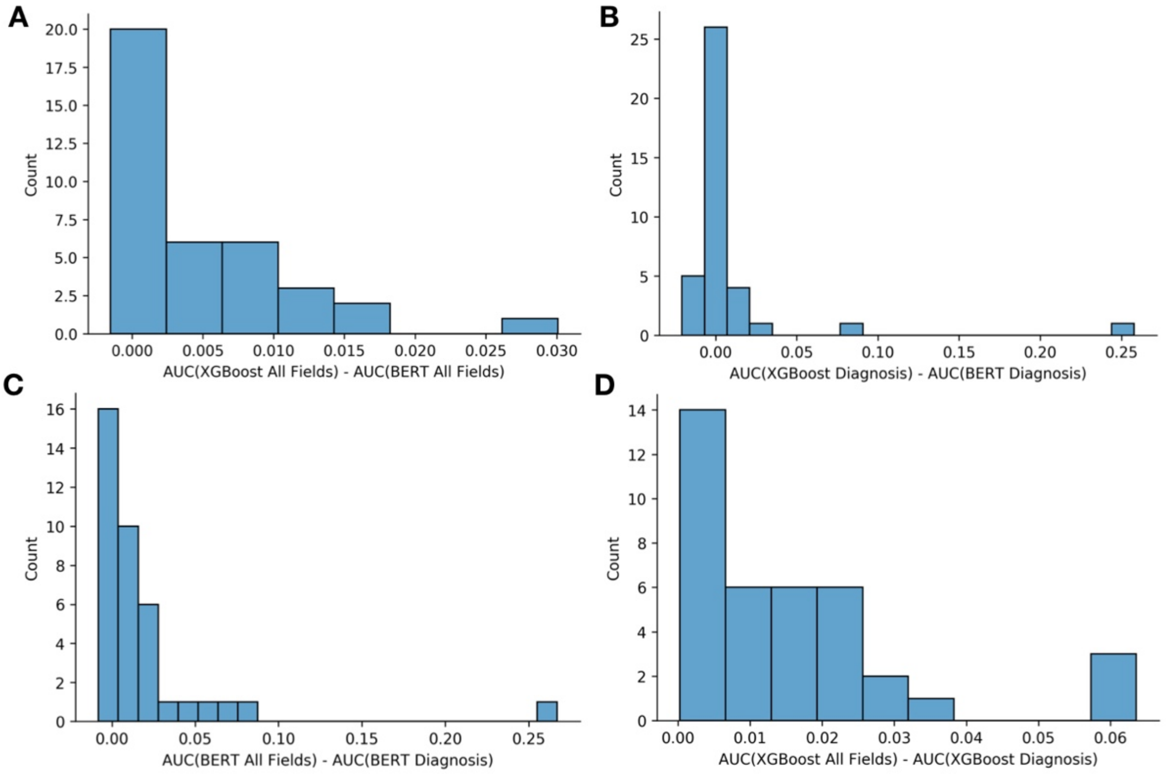

**Supplementary Figure 7:** Histogram of pairwise comparison (subtraction) of AUC statistics (averaged across cross-validation folds) between sets of algorithms / utilized document subfields; histogram tabulates AUC differences for individual codes, of which there are 38 values to be distributed amongst the histogram bins; reported relative performance gain (comparison/subtraction) of: **A)** XGBoost using all report subfields versus BERT using all report subfields, **B)** XGBoost using diagnostic subfield versus BERT using diagnostic subfield, **C)** BERT using all report subfields versus BERT using diagnostic subfield, **D)** XGBoost using all report subfields versus XGBoost using diagnostic subfield

**Supplementary Table 5:** Wilcoxon tests for significance of relative performance gains (distribution of paired AUC differences for codes between two algorithms/report subfield combinations); all Wilcoxon tests were one-sided (algorithm 1 / selected subfields performance greater than algorithm 2 / selected subfields performance) to see which models perform the best for CPT code prediction

| **Algorithm 1** | | **Algorithm 2** | | |
|---|---|---|---|---|
| **Name** | **Report Fields** | **Name** | **Report Fields** | **P-Value** |
| XGBoost | All Fields | BERT | All Fields | **2.8E-07** |
| XGBoost | Diagnosis | BERT | Diagnosis | 6.4E-01 |
| BERT | All Fields | BERT | Diagnosis | **4.2E-05** |
| XGBoost | All Fields | XGBoost | Diagnosis | **4.0E-08** |
| BERT | All Fields | SVM | All Fields | **4.0E-08** |
| BERT | Diagnosis | SVM | Diagnosis | **6.4E-05** |
| SVM | All Fields | SVM | Diagnosis | **6.8E-03** |

**Supplementary Table 6:** Sensitivity/Specificity for each algorithm/report subfield(s), averaged across cross-validation folds for each CPT code after optimization of Youden's index to select the sensitivity/specificity

| | BERT | | | | XGBoost | | | | SVM | | | |
|---|---|---|---|---|---|---|---|---|---|---|---|---|
| | Diagnosis | | All-Fields | | Diagnosis | | All-Fields | | Diagnosis | | All-Fields | |
| Code | Sensitivity | Specificity | Sensitivity | Specificity | Sensitivity | Specificity | Sensitivity | Specificity | Sensitivity | Specificity | Sensitivity | Specificity |
| 85060 | 1.00 | 1.00 | 1.00 | 1.00 | 1.00 | 1.00 | 1.00 | 1.00 | 1.00 | 1.00 | 1.00 | 1.00 |
| 85097 | 0.98 | 0.98 | 1.00 | 1.00 | 1.00 | 1.00 | 1.00 | 1.00 | 1.00 | 1.00 | 0.99 | 1.00 |
| 87491 | 0.96 | 0.98 | 0.99 | 1.00 | 0.99 | 0.98 | 1.00 | 1.00 | 0.99 | 0.98 | 0.99 | 0.97 |
| 87591 | 0.99 | 0.98 | 0.99 | 1.00 | 0.99 | 0.98 | 1.00 | 1.00 | 0.99 | 0.98 | 0.99 | 0.97 |
| 87624 | 0.98 | 0.99 | 0.98 | 0.99 | 0.98 | 0.99 | 0.98 | 0.99 | 0.98 | 0.97 | 0.97 | 0.98 |
| 88108 | 0.84 | 0.95 | 0.99 | 0.99 | 0.99 | 0.95 | 0.99 | 1.00 | 0.99 | 0.95 | 1.00 | 0.99 |
| 88112 | 0.97 | 0.96 | 0.99 | 0.99 | 0.99 | 0.97 | 1.00 | 0.99 | 0.99 | 0.97 | 0.99 | 0.99 |
| 88141 | 1.00 | 1.00 | 1.00 | 1.00 | 1.00 | 1.00 | 1.00 | 1.00 | 1.00 | 1.00 | 0.99 | 0.99 |
| 88142 | 0.95 | 0.89 | 0.99 | 0.97 | 0.99 | 0.93 | 0.97 | 0.97 | 0.99 | 0.93 | 0.94 | 0.95 |
| 88172 | 0.81 | 0.96 | 0.99 | 0.99 | 1.00 | 0.95 | 0.99 | 0.99 | 1.00 | 0.95 | 0.99 | 0.98 |
| 88173 | 1.00 | 0.97 | 1.00 | 0.99 | 1.00 | 0.97 | 1.00 | 0.99 | 0.99 | 0.97 | 0.99 | 0.99 |
| 88175 | 0.98 | 0.99 | 0.98 | 0.99 | 0.98 | 0.99 | 0.98 | 0.99 | 0.98 | 1.00 | 0.97 | 0.98 |
| 88177 | 0.83 | 0.95 | 1.00 | 0.99 | 0.99 | 0.95 | 1.00 | 1.00 | 0.98 | 0.95 | 1.00 | 0.99 |
| 88184 | 0.85 | 0.92 | 0.95 | 0.96 | 0.94 | 0.93 | 0.98 | 0.98 | 0.96 | 0.93 | 0.96 | 0.94 |
| 88185 | 0.71 | 0.90 | 0.95 | 0.95 | 0.94 | 0.93 | 0.98 | 0.97 | 0.96 | 0.92 | 0.95 | 0.95 |
| 88188 | 0.87 | 0.88 | 0.95 | 0.94 | 0.93 | 0.91 | 0.96 | 0.96 | 0.96 | 0.93 | 0.92 | 0.93 |
| 88189 | 0.77 | 0.73 | 0.94 | 0.95 | 0.88 | 0.88 | 0.95 | 0.97 | 0.95 | 0.95 | 0.92 | 0.95 |
| 88271 | 0.92 | 0.94 | 0.95 | 0.97 | 0.94 | 0.95 | 0.98 | 0.99 | 0.96 | 0.96 | 0.97 | 0.98 |
| 88274 | 0.96 | 0.94 | 0.97 | 0.99 | 0.95 | 0.95 | 0.99 | 0.99 | 0.97 | 0.97 | 0.98 | 0.99 |
| 88300 | 0.99 | 0.99 | 0.98 | 1.00 | 0.99 | 0.99 | 0.99 | 0.99 | 0.97 | 0.98 | 0.96 | 0.98 |
| 88302 | 0.90 | 0.98 | 0.94 | 0.96 | 0.96 | 0.97 | 0.96 | 0.97 | 0.95 | 0.93 | 0.93 | 0.92 |
| 88304 | 0.95 | 0.96 | 0.96 | 0.96 | 0.96 | 0.95 | 0.96 | 0.96 | 0.93 | 0.93 | 0.92 | 0.92 |
| 88305 | 0.94 | 0.90 | 0.96 | 0.92 | 0.93 | 0.91 | 0.95 | 0.95 | 0.20 | 0.68 | 0.18 | 0.70 |
| 88307 | 0.97 | 0.97 | 0.97 | 0.96 | 0.97 | 0.96 | 0.98 | 0.97 | 0.94 | 0.94 | 0.95 | 0.93 |
| 88309 | 0.96 | 0.97 | 0.96 | 0.97 | 0.97 | 0.97 | 0.98 | 0.98 | 0.94 | 0.95 | 0.96 | 0.95 |
| 88311 | 0.97 | 0.98 | 0.98 | 0.98 | 0.98 | 0.98 | 0.99 | 0.99 | 0.89 | 0.95 | 0.97 | 0.95 |
| 88312 | 0.84 | 0.88 | 0.93 | 0.94 | 0.93 | 0.92 | 0.98 | 0.98 | 0.85 | 0.86 | 0.84 | 0.84 |
| 88313 | 0.89 | 0.90 | 0.94 | 0.95 | 0.89 | 0.91 | 0.97 | 0.97 | 0.87 | 0.89 | 0.87 | 0.90 |
| 88321 | 0.98 | 0.95 | 0.99 | 0.99 | 0.96 | 0.95 | 1.00 | 1.00 | 0.91 | 0.91 | 0.99 | 0.99 |
| 88331 | 0.93 | 0.94 | 0.98 | 0.98 | 0.94 | 0.96 | 1.00 | 1.00 | 0.92 | 0.93 | 0.94 | 0.92 |
| 88332 | 0.92 | 0.93 | 0.96 | 0.95 | 0.94 | 0.94 | 0.99 | 0.99 | 0.87 | 0.95 | 0.95 | 0.96 |
| 88333 | 0.95 | 0.95 | 0.98 | 0.99 | 0.97 | 0.96 | 1.00 | 1.00 | 0.98 | 0.96 | 0.97 | 0.99 |
| 88341 | 0.84 | 0.83 | 0.91 | 0.91 | 0.85 | 0.85 | 0.96 | 0.95 | 0.80 | 0.80 | 0.90 | 0.89 |
| 88342 | 0.86 | 0.85 | 0.97 | 0.96 | 0.86 | 0.84 | 0.98 | 0.97 | 0.80 | 0.77 | 0.90 | 0.93 |
| 88344 | 0.95 | 0.97 | 0.96 | 0.97 | 0.94 | 0.94 | 0.97 | 0.98 | 0.95 | 0.97 | 0.94 | 0.98 |

| | | | | | | | | | | | | |
|---|---|---|---|---|---|---|---|---|---|---|---|---|
| 88346 | 0.92 | 0.91 | 0.99 | 0.99 | 0.99 | 1.00 | 1.00 | 1.00 | 0.98 | 0.97 | 0.98 | 1.00 |
| 88350 | 0.80 | 0.94 | 1.00 | 1.00 | 0.99 | 1.00 | 1.00 | 1.00 | 0.97 | 0.98 | 0.99 | 1.00 |
| 88360 | 0.91 | 0.93 | 0.95 | 0.96 | 0.93 | 0.93 | 0.97 | 0.97 | 0.92 | 0.94 | 0.94 | 0.94 |

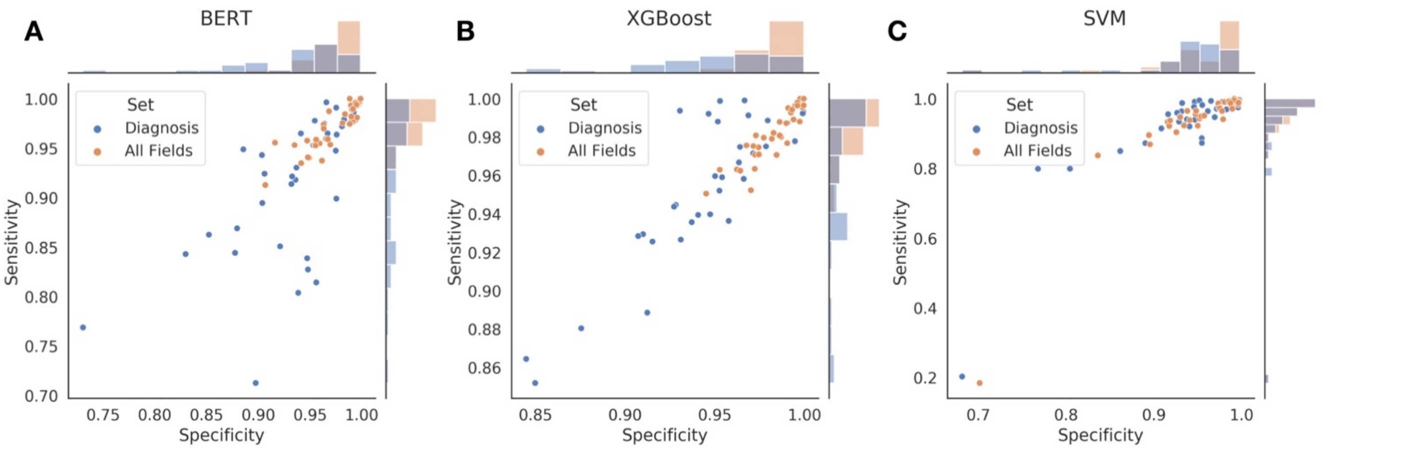

**Supplementary Figure 8:** Scatterplot of sensitivity and specificities for each CPT code, after averaging across CPT codes; individual point is a CPT code; point is colored by whether it was predicted from diagnostic text or all report subfields; histograms at plot margins indicate marginal distribution of code sensitivity/specificity

**Supplementary Table 7: First three numerical columns:** Averaged sensitivity and specificity across the XGBoost and BERT algorithms to denote overall predictive performance for each CPT code; Average Youden calculated from the sensitivity and specificity; **Final three numerical columns:** Changes in sensitivity, specificity and Youden when utilizing all report subfields versus the diagnostic text alone

| Code | Average Sensitivity | Average Specificity | Average Youden | Δ Sensitivity | Δ Specificity | Δ Youden |
|---|---|---|---|---|---|---|
| 85060 | 1.00 | 1.00 | 0.99 | 0.00 | 0.00 | 0.00 |
| 85097 | 0.99 | 0.99 | 0.99 | 0.01 | 0.01 | 0.02 |
| 87491 | 0.99 | 0.99 | 0.97 | 0.02 | 0.02 | 0.04 |
| 87591 | 0.99 | 0.99 | 0.98 | 0.00 | 0.02 | 0.02 |
| 87624 | 0.98 | 0.99 | 0.97 | 0.00 | 0.00 | 0.00 |
| 88108 | 0.95 | 0.97 | 0.92 | 0.08 | 0.04 | 0.12 |
| 88112 | 0.99 | 0.98 | 0.97 | 0.01 | 0.02 | 0.04 |
| 88141 | 1.00 | 1.00 | 1.00 | 0.00 | 0.00 | 0.00 |
| 88142 | 0.98 | 0.94 | 0.92 | 0.01 | 0.06 | 0.07 |
| 88172 | 0.95 | 0.97 | 0.92 | 0.08 | 0.04 | 0.12 |
| 88173 | 1.00 | 0.98 | 0.98 | 0.00 | 0.03 | 0.03 |
| 88175 | 0.98 | 0.99 | 0.97 | 0.00 | 0.00 | 0.00 |
| 88177 | 0.95 | 0.97 | 0.93 | 0.09 | 0.05 | 0.14 |
| 88184 | 0.93 | 0.95 | 0.88 | 0.07 | 0.04 | 0.11 |
| 88185 | 0.90 | 0.94 | 0.84 | 0.14 | 0.05 | 0.19 |

| | | | | | | |
|---|---|---|---|---|---|---|
| 88188 | 0.93 | 0.92 | 0.85 | 0.06 | 0.05 | 0.11 |
| 88189 | 0.89 | 0.88 | 0.77 | 0.12 | 0.16 | 0.28 |
| 88271 | 0.95 | 0.96 | 0.91 | 0.04 | 0.04 | 0.07 |
| 88274 | 0.97 | 0.97 | 0.94 | 0.02 | 0.04 | 0.06 |
| 88300 | 0.99 | 0.99 | 0.98 | 0.00 | 0.00 | 0.00 |
| 88302 | 0.94 | 0.97 | 0.91 | 0.02 | 0.00 | 0.02 |
| 88304 | 0.96 | 0.96 | 0.92 | 0.00 | 0.00 | 0.01 |
| 88305 | 0.94 | 0.92 | 0.86 | 0.02 | 0.03 | 0.04 |
| 88307 | 0.97 | 0.97 | 0.94 | 0.01 | 0.00 | 0.01 |
| 88309 | 0.97 | 0.97 | 0.94 | 0.00 | 0.00 | 0.01 |
| 88311 | 0.98 | 0.98 | 0.97 | 0.02 | 0.01 | 0.02 |
| 88312 | 0.92 | 0.93 | 0.85 | 0.07 | 0.07 | 0.14 |
| 88313 | 0.92 | 0.93 | 0.86 | 0.06 | 0.05 | 0.12 |
| 88321 | 0.98 | 0.97 | 0.96 | 0.03 | 0.04 | 0.07 |
| 88331 | 0.96 | 0.97 | 0.93 | 0.05 | 0.04 | 0.09 |
| 88332 | 0.95 | 0.95 | 0.90 | 0.04 | 0.03 | 0.07 |
| 88333 | 0.98 | 0.98 | 0.95 | 0.03 | 0.03 | 0.06 |
| 88341 | 0.89 | 0.88 | 0.78 | 0.09 | 0.09 | 0.18 |
| 88342 | 0.92 | 0.91 | 0.83 | 0.11 | 0.12 | 0.23 |
| 88344 | 0.95 | 0.97 | 0.92 | 0.02 | 0.02 | 0.04 |
| 88346 | 0.98 | 0.97 | 0.95 | 0.03 | 0.04 | 0.08 |
| 88350 | 0.95 | 0.98 | 0.93 | 0.10 | 0.03 | 0.13 |
| 88360 | 0.94 | 0.95 | 0.89 | 0.04 | 0.03 | 0.08 |

**Supplementary Table 8:** Classification reports for pathologist prediction models (BERT, XGBoost, SVM) for reported subfields (diagnostic/all-fields)

| BERT | | | | | | | |
|---|---|---|---|---|---|---|---|
| Diagnosis | | | | All-Fields | | | |
| Pathologist | Precision | Recall | F1-Score | Pathologist | Precision | Recall | F1-Score |
| 1 | 0.94 | 0.94 | 0.94 | 1 | 0.95 | 0.94 | 0.94 |
| 2 | 0.49 | 0.82 | 0.61 | 2 | 0.61 | 0.84 | 0.70 |
| 3 | 0.94 | 0.86 | 0.89 | 3 | 0.99 | 0.98 | 0.98 |
| 4 | 0.77 | 0.76 | 0.77 | 4 | 0.81 | 0.81 | 0.81 |
| 5 | 0.80 | 0.85 | 0.82 | 5 | 0.88 | 0.88 | 0.88 |
| 6 | 0.93 | 0.98 | 0.95 | 6 | 0.96 | 0.96 | 0.96 |
| 7 | 0.81 | 0.82 | 0.81 | 7 | 0.87 | 0.87 | 0.87 |
| 8 | 0.36 | 0.91 | 0.51 | 8 | 0.41 | 0.80 | 0.55 |
| 9 | 0.86 | 0.78 | 0.82 | 9 | 0.86 | 0.80 | 0.83 |
| 10 | 0.78 | 0.61 | 0.69 | 10 | 0.74 | 0.68 | 0.71 |
| 11 | 0.67 | 0.71 | 0.69 | 11 | 0.71 | 0.73 | 0.72 |
| 12 | 0.84 | 0.77 | 0.80 | 12 | 0.87 | 0.83 | 0.85 |
| 13 | 0.80 | 0.91 | 0.85 | 13 | 0.86 | 0.91 | 0.88 |
| 14 | 0.72 | 0.74 | 0.73 | 14 | 0.83 | 0.85 | 0.84 |
| 15 | 0.83 | 0.74 | 0.78 | 15 | 0.84 | 0.83 | 0.83 |
| 16 | 0.56 | 0.25 | 0.34 | 16 | 0.54 | 0.35 | 0.42 |
| 17 | 0.89 | 0.96 | 0.93 | 17 | 0.93 | 0.96 | 0.94 |
| 18 | 0.58 | 0.14 | 0.22 | 18 | 0.45 | 0.27 | 0.34 |
| 19 | 0.71 | 0.72 | 0.71 | 19 | 0.71 | 0.74 | 0.72 |
| 20 | 0.84 | 0.39 | 0.53 | 20 | 0.74 | 0.43 | 0.54 |
| Accuracy | 0.74 | 0.74 | 0.74 | Accuracy | 0.79 | 0.79 | 0.79 |
| Macro Avg | 0.76 | 0.73 | 0.72 | Macro Avg | 0.78 | 0.77 | 0.77 |
| Weighted Avg | 0.77 | 0.74 | 0.74 | Weighted Avg | 0.80 | 0.79 | 0.79 |

**XGBoost**

| Diagnosis | | | | All-Fields | | | |
|---|---|---|---|---|---|---|---|
| Pathologist | Precision | Recall | F1-Score | Pathologist | Precision | Recall | F1-Score |
| 1 | 0.92 | 0.88 | 0.90 | 1 | 0.94 | 0.89 | 0.91 |
| 2 | 0.67 | 0.66 | 0.67 | 2 | 0.68 | 0.76 | 0.72 |
| 3 | 0.90 | 0.85 | 0.88 | 3 | 1.00 | 1.00 | 1.00 |
| 4 | 0.81 | 0.76 | 0.78 | 4 | 0.80 | 0.83 | 0.81 |
| 5 | 0.74 | 0.89 | 0.81 | 5 | 0.86 | 0.91 | 0.88 |
| 6 | 0.94 | 0.98 | 0.96 | 6 | 0.97 | 0.98 | 0.97 |
| 7 | 0.88 | 0.77 | 0.82 | 7 | 0.92 | 0.88 | 0.90 |
| 8 | 0.36 | 0.87 | 0.51 | 8 | 0.51 | 0.73 | 0.60 |
| 9 | 0.72 | 0.88 | 0.79 | 9 | 0.79 | 0.86 | 0.82 |
| 10 | 0.80 | 0.62 | 0.70 | 10 | 0.76 | 0.67 | 0.72 |
| 11 | 0.75 | 0.77 | 0.76 | 11 | 0.78 | 0.76 | 0.77 |
| 12 | 0.73 | 0.81 | 0.77 | 12 | 0.79 | 0.87 | 0.83 |
| 13 | 0.83 | 0.76 | 0.79 | 13 | 0.92 | 0.87 | 0.90 |
| 14 | 0.78 | 0.68 | 0.73 | 14 | 0.91 | 0.83 | 0.87 |
| 15 | 0.75 | 0.73 | 0.74 | 15 | 0.83 | 0.82 | 0.82 |
| 16 | 0.50 | 0.32 | 0.39 | 16 | 0.56 | 0.47 | 0.51 |
| 17 | 0.69 | 0.52 | 0.59 | 17 | 0.88 | 0.76 | 0.81 |
| 18 | 0.69 | 0.21 | 0.32 | 18 | 0.58 | 0.42 | 0.48 |
| 19 | 0.71 | 0.74 | 0.72 | 19 | 0.71 | 0.75 | 0.73 |
| 20 | 0.83 | 0.38 | 0.53 | 20 | 0.70 | 0.51 | 0.59 |
| Accuracy | 0.73 | 0.73 | 0.73 | Accuracy | 0.80 | 0.80 | 0.80 |
| Macro Avg | 0.75 | 0.70 | 0.71 | Macro Avg | 0.79 | 0.78 | 0.78 |
| Weighted Avg | 0.75 | 0.73 | 0.72 | Weighted Avg | 0.80 | 0.80 | 0.80 |

**SVM**

| Diagnosis | | | | All-Fields | | | |
|---|---|---|---|---|---|---|---|
| Pathologist | Precision | Recall | F1-Score | Pathologist | Precision | Recall | F1-Score |
| 1 | 0.59 | 0.62 | 0.60 | 1 | 0.45 | 0.50 | 0.47 |
| 2 | 0.38 | 0.36 | 0.37 | 2 | 0.10 | 0.00 | 0.00 |
| 3 | 0.56 | 0.57 | 0.56 | 3 | 0.86 | 0.84 | 0.85 |
| 4 | 0.33 | 0.16 | 0.22 | 4 | 0.20 | 0.18 | 0.19 |
| 5 | 0.39 | 0.52 | 0.44 | 5 | 0.24 | 0.73 | 0.36 |
| 6 | 0.36 | 0.56 | 0.44 | 6 | 0.34 | 0.65 | 0.45 |
| 7 | 0.09 | 0.04 | 0.05 | 7 | 0.00 | 0.00 | 0.00 |
| 8 | 0.36 | 0.80 | 0.49 | 8 | 0.34 | 0.92 | 0.49 |
| 9 | 0.49 | 0.67 | 0.57 | 9 | 0.28 | 0.79 | 0.41 |
| 10 | 0.34 | 0.09 | 0.14 | 10 | 0.18 | 0.05 | 0.07 |
| 11 | 0.44 | 0.32 | 0.38 | 11 | 0.23 | 0.32 | 0.26 |
| 12 | 0.24 | 0.36 | 0.29 | 12 | 0.21 | 0.24 | 0.23 |
| 13 | 0.00 | 0.00 | 0.00 | 13 | 0.00 | 0.00 | 0.00 |
| 14 | 0.00 | 0.00 | 0.00 | 14 | 0.00 | 0.00 | 0.00 |
| 15 | 0.23 | 0.42 | 0.30 | 15 | 0.26 | 0.11 | 0.16 |
| 16 | 0.30 | 0.18 | 0.23 | 16 | 0.28 | 0.04 | 0.07 |
| 17 | 0.00 | 0.00 | 0.00 | 17 | 0.00 | 0.00 | 0.00 |
| 18 | 0.06 | 0.02 | 0.02 | 18 | 0.18 | 0.03 | 0.05 |
| 19 | 0.32 | 0.49 | 0.38 | 19 | 0.00 | 0.00 | 0.00 |
| 20 | 0.07 | 0.02 | 0.03 | 20 | 0.11 | 0.00 | 0.00 |
| Accuracy | 0.35 | 0.35 | 0.35 | Accuracy | 0.32 | 0.32 | 0.32 |
| Macro Avg | 0.28 | 0.31 | 0.28 | Macro Avg | 0.21 | 0.27 | 0.20 |
| Weighted Avg | 0.29 | 0.35 | 0.30 | Weighted Avg | 0.24 | 0.32 | 0.24 |

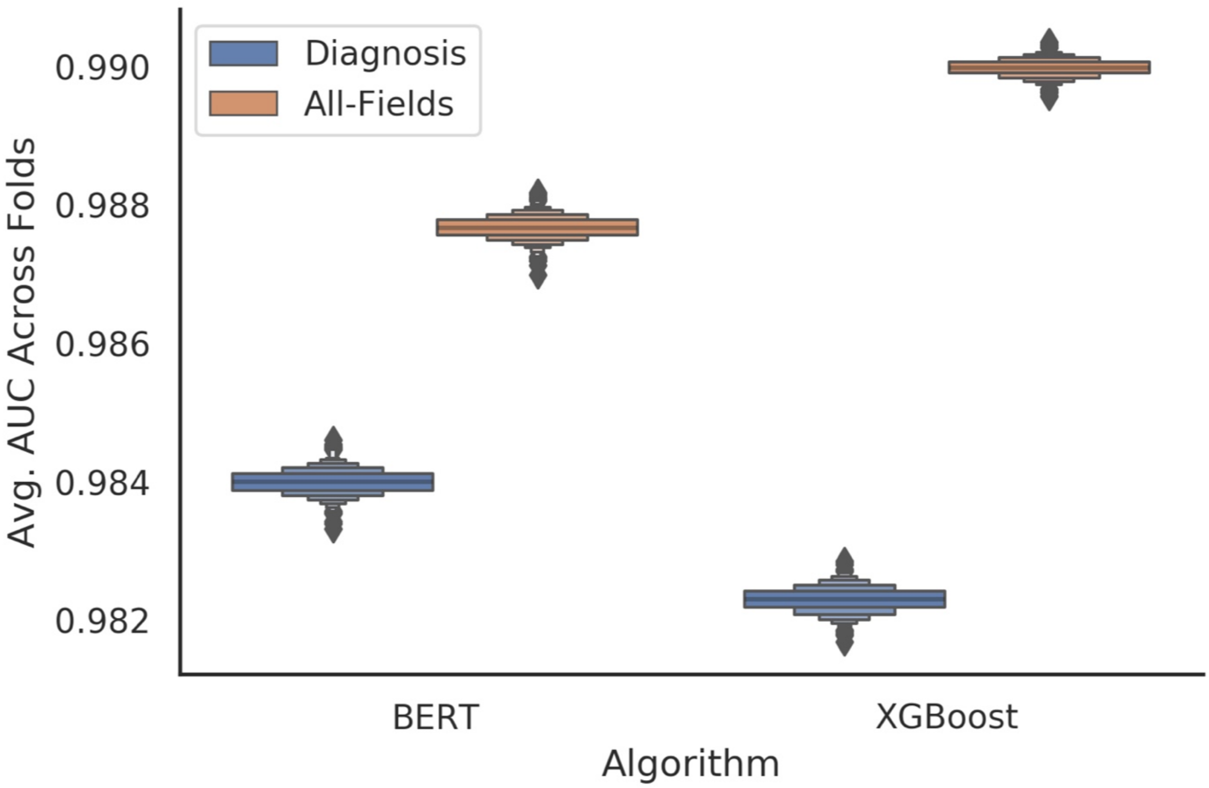

**Supplementary Figure 9**: Averaged weighted AUC statistics across pathologists/cross-validation folds for prediction of top 20 pathologists with most sign-outs; reports for BERT and XGBoost for the diagnosis and all-fields models

**Additional Description of Topic Modeling and Report Characterization Techniques: TF-IDF, UMAP, HSBSCAN, LDA**

Here, we briefly provide an overview of the modeling techniques that, when utilized in conjunction, characterized the pathology report corpus through the establishment of important words that were not ubiquitous across the corpus (TF-IDF), removal of noise and discovery of clusters (UMAP and HDBSCAN) and generating topics which describe recurrent themes (LDA).

TF-IDF (term frequency inverse document frequency) takes as input a sparse count matrix, which contains the reports as rows and individual words/n-grams as columns, where each element is a count of the n-gram in the document. TF-IDF re-weights the count matrix based on an algorithm that modifies word importance on the basis of whether the word is ubiquitous across all documents and/or enriched in its own document. The formula for TF-IDF is:

$$tf - idf_{t,d} = tf_{t,d} * \log\frac{n}{1+\mathrm{df}_t} + 1$$

Where $t$ and $d$ refer to the specific term and document respectively. The term-frequency, $tf$, is the reported count of the n-gram in the particular document, while document frequency, $df$, is the number of reports that contain the term (i.e. how ubiquitous the word is across the corpus). Normally these values are normalized via the euclidean norm to downweight longer documents.

Such information may replace the count matrix for downstream analysis, though is not necessary.

UMAP operates on the count/tf-idf matrix to reduce the dimensionality of the reports while preserving the key relationships between the reports. This is unlike PCA, which selects principal components to maximize variance, and TSNE (T-Stochastic Neighborhood Embedding), which learns a lower dimensional manifold that preserves local distance between reports. UMAP (Uniform Manifold Approximation and Projection for Dimension Reduction) forms fuzzy simplicial sets that represents the higher dimensional manifold at multiple distances. Computationally, this amounts to constructing a weighted nearest neighbors graph and an optimization routine that preserves similar structure in the low dimensional manifold while optimizing a force-directed graph layout.

HDBSCAN is a clustering algorithm, which operates on the lower dimensional manifold to find natural groupings of the data. HDBSCAN combines hierarchical clustering techniques, which iteratively merges similar clusters, with density-based clustering, which estimates clusters of similar density. HDBSCAN estimates the density of points based on whether a certain number of points exist within a small well-defined neighborhood and whether two points share a common neighbor, both outside of that which is expected if there were noise. HDBSCAN varies the size of this neighborhood to consider/integrate density on multiple scales to form a hierarchy, which may be further processed to yield the clusters. This yields a set of clusters and points that have been defined as noise. Since the algorithm considers the notion of distance and connectedness on multiple scales / neighborhoods, it often pairs well with UMAP due to similarities in formulation.

Latent Dirichlet Allocation (LDA) is a 3-level probabilistic/Bayesian generative model for inferring a distribution of topics across a document corpus. Ultimately, the goal of the model is to provide a mechanistic model for how the count matrix arises (that is, estimating the frequency of words in each of the reports). The simplified conceptual framework is as follows:

1. A document is selected.
2. $N$ number of words are selected from a Poisson distribution (which iterates steps 3-4 $N$ times).
3. For each word (not yet selected), a topic is selected from a set of latent topics (topic mixture) that characterize the document with some probability.
4. A word is selected from a set of words that are ascribed to the topic.
5. The distribution of words selected via the generative approach in steps 1-3 are compared to the true distribution of words after marginalizing over the topics, and documents.

The generative model initially places two separate Dirichlet priors over selecting topics (topic mixture) and words from topics (a k words by V topics matrix). Variational bayes and expectation maximization techniques are applied to estimate the posterior distribution of the topic mixture and topic-word parameters by assuming the conditional posterior follows a known family of distributions. Ultimately, sampling the predictive posterior allows for inference of the distribution of topics across documents.

**Additional Description of Explanation Techniques: SHAP, Integrated Gradients, Self-Attention, Attention Over Pathology Report Subfields**

Shapley additive feature explanations (SHAP) is a technique that explains the results of any machine learning model, which may have a complex decision surface. SHAP approximates this surface on a sample-by-sample basis by fitting one local additive model per sample. The coefficients of this model represent the importance of a feature or word. Local additive models operate to estimate directly estimate the prediction of the machine learning model when summed. That is, if $f$ is the machine learning model, $g$, for pathology report $i$, with term frequency $x_i$, then the approximation is as follows:

$$f(x_i) \approx g_i(x_i) = E[f(z)] + \sum_k \phi_{k,i}(f, x_i)$$

Here, $\phi_{k,i}$ represents the shapley coefficient for term $k$ of report $i$. The fitting procedure decides how to distribute the remainder between the mean value of the learned model over the dataset and the prediction to each of the predictors, while considering the importance of the individual predictor over the permutation or ensemble of possible orderings of predictors when assigning reward (remainder). The predictor importances derived for individual CPT codes or pathologists were estimated by averaging these term/word level importances / shapley coefficients across the entire document (we subsampled with a random seed for a more efficient computation) report corpus for a given model. This analysis was conducted for the XGBoost modeling approaches.

We utilized integrated gradients to interpret which words were found to be important for individual sentences when utilizing the BERT model, which we applied on the diagnostic text. Integrated gradients is a backpropagation based method for identifying salient features. Many traditional methods for ascertaining important predictors will take the gradient of the model prediction with a defined input $\nabla F(\vec{x})$, which serves as a linear approximation to the complex functional approximation, and multiply by the original input, $\vec{x}$, to yield the predictor specific importance ($\vec{x} \odot \nabla F(\vec{x})$). However, this is less than ideal when $\vec{x}$ exists in the domain where the gradient saturates and also has no baseline for comparison. Integrated gradients, related to shapley values, overcomes these two issues by first establishing a noninformative baseline/counterfactual $\overrightarrow{x_0}$, then successively sums more informative gradients along the path from the baseline to the observation $\overrightarrow{x_i}$ to yield the overall importance of the predictors:

$$IG(\overrightarrow{x_i}) = (\overrightarrow{x_i} - \overrightarrow{x_0}) * \int_{\alpha=0}^{\alpha=1} \nabla F\left(\overrightarrow{x_0} + \alpha(\overrightarrow{x_i} - \overrightarrow{x_0})\right)$$

Much of the success of the BERT methodology can be attributed to a neural network modeling approach known as the Transformer. As input to the model, each word is mapped to a semantic vector that captures the word's meaning, which is updated throughout the training process. The Transformer contextualizes the set of word vectors in a report through its encoder and decoder layers. The encoder and decoder layers are further decomposed into self-attention and feed-forward neural networks. Self-attention mechanisms capture dependencies between words within the sentence by forming a weight between each word and individually all of the words of the sentence; that is, identifying the most relevant words for the understanding of the current word. This is accomplished by estimating a weight between two words of a sentence. Here, matrix operations may be employed to speed up the calculation of the self-attention.

Suppose the word embeddings of the sentence are encapsulated in matrix $\overleftrightarrow{X}$, where rows indicate words and columns are the latent dimensions. Parameterized query, key and value matrices are generated via the following operations:

$$\overleftrightarrow{Q} = W_Q\overleftrightarrow{X}$$
$$\overleftrightarrow{K} = W_K\overleftrightarrow{X}$$
$$\overleftrightarrow{V} = W_V\overleftrightarrow{X}$$

The query and key vectors are utilized as follows to construct the paired attention weights across a sentence, which could be thought of as learning/estimating a weighted unipartite matrix, attention matrix $\overleftrightarrow{A}$ ($d_k$ is used for further normalization):

$$\overleftrightarrow{A} = softmax\left(\frac{\overleftrightarrow{Q}\overleftrightarrow{K}^T}{\sqrt{d_k}}\right)$$

$\overleftrightarrow{A}$ is the estimate of the word-to-word dependencies in the sentence for this particular operation. The embeddings of the sentence are updated/contextualized by multiplying this self-attention matrix with the embedding values:

$$\overleftrightarrow{Z} = \overleftrightarrow{A}\overleftrightarrow{V}$$

Usually, these self-attention matrices represent particular dependencies within the sentence. However, there may exist many complex dependencies to build a global understanding of the sentence/paragraph/report. As such, multiple self-attention "heads" are generated by allowing the existence of many query, key and value matrices per encoding layer. We visualized the output of the estimated self-attention matrices in our manuscript to demonstrate some of the learned dependencies. We have also omitted from this discussion nuanced specifics pertaining to the decoder (eg. retaining the query and key matrices from the encoder layers), residual connections and positional embeddings, as they do not necessarily pertain to methods to interpret the output of the BERT model for a pathology report.

Attention across document subfields is entirely separate from BERT self-attention mechanisms. As mentioned in the main text, attention weights are utilized to decide how much information from report subsections to incorporate into the final global representation of the report. A weight matrix $W$, an $n_z$ (number of latent dimensions) by one matrix, serves as a filter/gate to score how important a subsection is. The scores for each of the report subfields are softmaxed to assign a probability to each subfield for incorporation, $\vec{\alpha}$. The gate is learned via model parameter updates during backpropagation. Importantly, we report the attention weights $\vec{\alpha}$ to communicate the importance of specific report subsections.
